# Airborne transmission of SARS-CoV-2 over distances greater than two metres: a rapid systematic review

**DOI:** 10.1101/2021.10.19.21265208

**Authors:** Jennifer C Palmer, Daphne Duval, Isobel Tudge, Jason Kwasi Sarfo-Annin, Nicola Pearce-Smith, Emer O’Connell, Allan Bennett, Rachel Clark

**Affiliations:** COVID-19 Rapid Evidence Service, UK Health Security Agency (UKHSA), UK; MRC Integrative Epidemiology Unit at the University of Bristol, UK; Population Health Science, Bristol Medical School, University of Bristol, UK; COVID-19 Advice and Guidance, UK Health Security Agency (UKHSA), UK; Research and Evaluation, UK Health Security Agency (UKHSA), UK

## Abstract

**Objective:** To evaluate the potential for long-distance (over two metres) airborne transmission of SARS-CoV-2 in indoor community settings and investigate factors which may impact this transmission.

**Design:** Systematic review and narrative synthesis.

**Data source:** MEDLINE, Embase, medRxiv, Arxiv and WHO COVID-19 Research Database for studies published from 27 July 2020 to 21 April 2021; existing relevant rapid systematic reviews for studies published between 1 January to 27 July 2020.

**Eligibility criteria for study selection:** Observational studies that included a thorough epidemiological assessment of routes of transmission and which reported on the likelihood of airborne transmission of SARS-CoV-2 at a distance greater than two metres in indoor community settings.

**Data extraction and synthesis:** Data extraction was completed by one reviewer and independently checked by a second reviewer. Primary outcomes were COVID-19 infections via airborne transmission over distances greater than two metres and any factors that may have modified transmission risk. Included studies were rated using a quality criteria checklist (QCC) for primary research and certainty of key outcomes was determined using GRADE. Narrative synthesis was themed by setting.

**Results:** Of the 3,780 articles screened for inclusion, 15 publications reporting on 13 epidemiological investigations were included (three high, six medium and four low quality). Airborne transmission at distances greater than two metres was likely to have occurred for some or all transmission events in 12 studies and was unclear in one study (GRADE: very low certainty). In all studies, one or more factors plausibly increased the likelihood of long-distance airborne transmission occurring, particularly insufficient air replacement (GRADE: very low certainty), recirculating air flow (GRADE: very low certainty) and singing (GRADE: very low certainty). In nine studies, the primary cases were reported as being asymptomatic, presymptomatic or around symptom onset at the time of transmission.

**Conclusion:** This rapid systematic review found evidence of long-distance airborne transmission of SARS-CoV-2 in indoor community settings and identified factors that likely contributed to this transmission in all included studies. These results strengthen the need for adequate mitigation measures in indoor community settings, particularly adequate ventilation with fresh air, and caution required with the use of recirculating air flow systems.

**Systematic review registration:** PROSPERO CRD42021236762

## INTRODUCTION

COVID-19 is a viral infection transmitted through respiratory particles that contain SARS-CoV-2 virus. Since the start of the pandemic, direct transmission via respiratory particles with ballistic trajectory that directly deposit on mucous membranes, usually within two metres, has been considered the main transmission route for SARS-CoV-2. Transmission via fomites is another route, but this is considered to be relatively minimal by many experts.^1^ However, the contribution of airborne transmission via respiratory particles that remain suspended in the air for extended periods of time and are subsequently inhaled is less clear.^2-4^ The epidemiological challenge has been complicated by differences in terminologies, definitions and size thresholds for different respiratory particles which vary across disciplines. Regardless of terminology, respiratory particles are those that can be inhaled directly from the air, which is more likely to happen at short-range where the concentration of particles is higher than over a longer distance.^2, 4^

Distance and risk of transmission of SARS-CoV-2 are important considerations for risk management and public health decision making, especially in relation to infection prevention and control measures. Whether through inhalation of particles suspended in the air or through direct contact with ballistic particles, close contact (less than two metres) is accepted as having the highest risk for transmission, and is the rationale behind the recommendation in England to maintain a distance of at least two metres between people to reduce transmission.^5^ Outdoors, the risk of transmission of SARS-CoV-2 at distances greater than two metres is considered to be low due to dilution, dispersion and decay.^6, 7^ However, the evidence for transmission beyond two metres is still uncertain for indoor settings, where the concentration of respiratory particles can remain suspended in the air for longer and build-up, particularly in poorly-ventilated spaces, potentially resulting in an increased long-range transmission risk. As new, more transmissible variants of SARS-CoV-2 emerge,^8^ it is important to understand if transmission occurs at distances greater than two metres, and if it does, what factors may impact this risk.

A previous review that included 14 studies, mainly environmental sampling studies in healthcare settings and modelling studies, concluded that airborne transmission of SARS-CoV-2 in indoor settings was possible.^9^ Since then, the body of evidence on airborne transmission of SARS-CoV-2 has grown and more recent reviews were inconclusive on the nature and impact of airborne transmission of SARS-CoV-2.^10, 11^ These reviews include more studies with a wide range of study designs including observational (mainly epidemiological), experimental (such as environmental sampling) and modelling (such as fluid dynamic simulation) studies and also include studies from community settings. Few air sampling studies have detected viable virus (by viral culture of samples); instead most rely only on detection of SARS-CoV-2 viral RNA by RT-PCR, which does not distinguish between live virus, dead virus or RNA fragments. Additionally, many of these sampling studies were undertaken in acute hospital environments, where many cases will be past the period of peak shedding due to the lag in presentation of severe disease, which may underestimate infectiousness.

It is proposed that epidemiological studies, that can be considered as strong evidence for biological plausibility of airborne transmission,^12^ are the best evidence currently available to assess where and when transmission likely occurred. To enable this assessment, epidemiological investigations need to be extensive enough to allow a distinction of the most likely transmission routes and to assess whether airborne transmission at distances greater than two metres is likely to have occurred. There is a need to systematically identify and examine such studies to evaluate the potential for long-range airborne transmission of SARS-CoV-2 in indoor community settings and to assess the impact of potential transmission modifying factors.

## METHODS

A rapid systematic review approach was employed, following streamlined systematic methodologies to accelerate the review process,^13^ and reported according to PRISMA guidelines.^14^ The protocol for this review was registered on PROSPERO prior to screening (2021 CRD42021236762).^15^

### Data sources and searches

Primary studies were identified through two different sources. For studies published between 1 January 2020 and 27 July 2020 the included studies from a rapid systematic review by Comber et al^10^ were screened. This systematic review, assessed to be of moderate quality using the AMSTAR 2 critical appraisal tool,^16^ contains a comprehensive search strategy and a wider inclusion criteria than this rapid review (studies related to all airborne transmission of SARS-CoV-2, regardless of distance) and was the only relevant review available at the time of writing this protocol.

For search dates from 27 July 2020 to 21 April 2021, electronic searches were conducted in MEDLINE, Embase, medRxiv, Arxiv and WHO COVID-19 Research Database (initial search conducted on the 8 February 2021 and updated on the 21 April 2021). The full search strategy is provided in Supplementary Fiigure 1.

### Eligibility criteria for study selection

Published articles, accepted manuscripts and preprints reporting on potential for airborne transmission of SARS-CoV-2 in indoor community settings at a distance greater than approximately two metres (2m threshold is based on UK regulations were included along with non-UK-based studies that used other thresholds according to their respective national recommendations such as 1.5m or 6 feet/1.8m). The aim was to include all observational studies (outbreak investigations and epidemiological case series, cohort, case-control and cross-sectional studies) of any human population in community settings. Systematic or narrative reviews, guidelines, opinion pieces, intervention studies, modelling studies, environmental sampling studies without epidemiological investigation, laboratory or virology studies and animal studies were excluded. Observational studies where transmission via other routes such as close contact transmission or fomite transmission were likely to have been the main transmission route were also excluded (for example, studies reporting on transmission within households were excluded).

Screening was performed in Rayyan. Titles and abstracts of the first 10% of retrieved records were independently screened by two reviewers (97% agreement based on Cohen’s kappa coefficient). The remainder were screened by a single reviewer with a further 10% selected at random for screening by a second reviewer (99% agreement based on Cohen’s kappa coefficient). All records selected were screened at full text by one reviewer and checked for agreement by a second. Any discrepancies were resolved by discussion between the two reviewers.

### Outcomes

The primary outcomes were COVID-19 infections via airborne transmission at a distance greater than two metres, and any factors that might have modified transmission risk (‘modifying factors’). Examples of modifying factors that could increase risk of airborne transmission at a distance greater than two metres includes insufficient air replacement (such as no door or window opened) and recirculating air flow (for instance through mechanical ventilation that can generate air currents). Additional outcomes extracted, where available, were time spent in the setting and distance over which airborne transmission was thought to have occurred.

### Data extraction and synthesis

A data extraction table was developed to gather information on methods, participants, settings and key findings. Data extraction was completed for each included study by one reviewer, independently checked by a second reviewer and any discrepancies were resolved via discussion. Only evidence directly relevant to the review question was extracted. For instance, results on clinical outcomes were not extracted. Similarly, for studies that reported on different outbreaks or on onward transmission that might have happened in different settings, only the results of outbreaks or settings where distance and transmission routes could be assessed were extracted.

A narrative summary of results according to indoor setting is presented.

### Quality assessment and certainty of evidence

A quality criteria checklist (QCC) for primary research was used to assess the methodological quality of each included study.^17^ This tool can be applied to most study designs and was therefore considered suitable for rapid reviews of mixed types of evidence. The QCC tool is composed of 10 validity questions based on quality criteria and domains identified by the Agency for Healthcare Research and Quality (AHRQ),^18^ and four out of the 10 validity questions are considered critical (on selection bias, group comparability, description of exposure/assessment of transmission routes, and validity of outcome measurements). Strict criteria were used to assess the two critical questions related to exposure and outcome assessment. In particular, cluster of cases in the setting of interest had to be confirmed with viral genomic sequencing to be considered as low risk of bias for validity of outcome measurements.

A study is rated as high quality if the answers to the four critical questions are ‘yes’ plus at least one of the remaining questions (allowing one ‘unclear’ for a critical question if all remaining questions answer ‘yes’). The study is rated as low quality if ≥50% of the critical questions answered ‘no’ and/or greater than 50% of the remaining questions answer ‘no’. Otherwise, the study is rated as medium quality. Each study was assessed independently in duplicate and disagreements were resolved by consensus.

The certainty of the evidence was assessed using a variation of the GRADE framework for systematic reviews without meta-analysis.^19^ Each of the five GRADE domains (methodological limitations of the studies, indirectness, imprecision, inconsistency and the likelihood of publication bias) was assessed and classified as ‘no limitation or not serious’ (not important enough to warrant downgrading), ‘serious’ (downgrading the certainty rating by one level) or ‘very serious’ (downgrading the certainty rating by two levels). The body of evidence for a specific outcome was then classified as high certainty, moderate certainty, low certainty or very low certainty.

## RESULTS

### Study selection

The PRISMA flow diagram is shown in Figure 1. Following removal of duplicates, 3,780 records were screened for relevance using titles and abstracts, with 69 full texts assessed for eligibility. A further 54 studies were excluded at this stage (see Supplementary Tabble 1 for a list of excluded studies with reasons for exclusion). Fifteen publications^20-34^ reporting on 13 studies were included in this review. When two publications reported on the same study, only the main publication (the most comprehensive one) was extracted and rated for quality.

**Figure 1.**
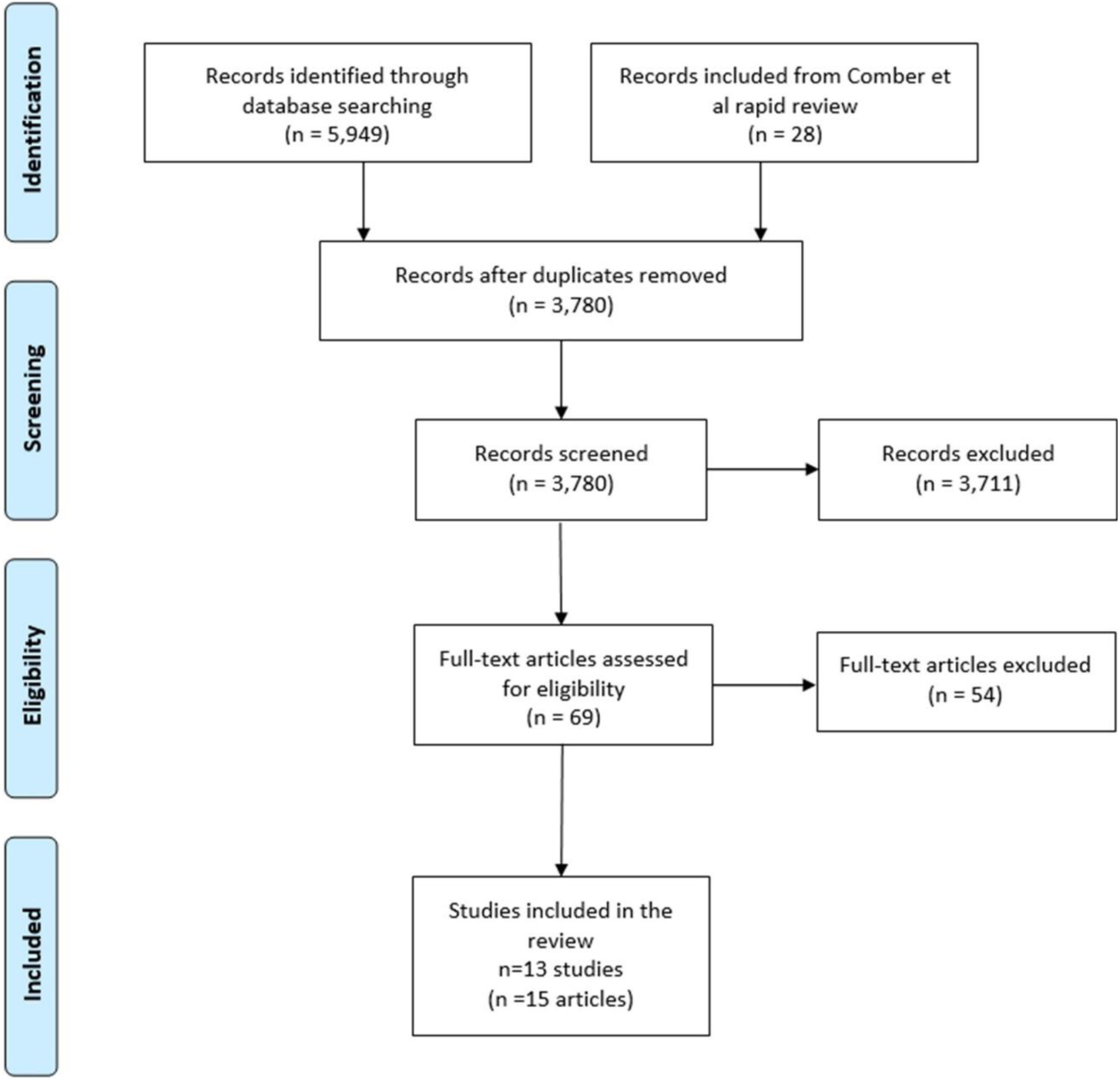
PRISMA flow diagram.

All studies were epidemiological outbreak investigations of community clusters of COVID-19 infections, one of which also contains a nested case-control study.^32^ Out of the 13 studies, seven were conducted in Asia,^24, 25, 27-30, 32^ three in Europe,^20, 22, 31^ two in Oceania^21, 26^ and one in the USA.^23^ In terms of settings, two studies reported on transmission in restaurants,^27, 28^ two between apartments in an apartment block,^24, 29^ two in buses,^30, 32^ one in a food processing factory,^22^ one in a quarantine hotel,^21^ one in a department store^25^ and four during singing events.^20, 23, 26, 31^

Details of the included studies are provided in Supplementary Tabble 2 and a summary of each study in Table 1.

**Table 1.** Summary of included studies and outcome assessment.

| Reference | Study setting and period | Number of COVID-19 cases | Outcome and exposure assessment | Potential for other transmission routes* | Potential for airborne transmission > 2m*†‡ | Modifying factors |
| --- | --- | --- | --- | --- | --- | --- |
| Charlotte et al, 2020<sup>20</sup><br><u>Quality rating:</u><br>low | Singing event, France<br>March 2020 | 19 COVID-19 cases (7 confirmed, 12 probable) | No genomic sequencing<br>Questionnaire and telephone interviews | Close contact: possible for some cases<br>Fomite transmission: unlikely<br>Transmission from outside of event: probable for at least 2 cases | **Possible** airborne transmission >2m for some cases, due to high SAR (2h) | Insufficient air replacement<br>Singing |
| Eichler et al, 2021<sup>21</sup><br><u>Quality rating:</u><br>low | Quarantine hotel, New Zealand<br>September 2020 | 9 confirmed cases, but only 1 considered for long-distance transmission | Genomic sequencing<br>Epidemiologic data, surveillance video, review of the ventilation system in the hotel | Close contact: unclear<br>Fomite transmission: unclear<br>Transmission from outside of event: unlikely | **Possible** airborne transmission from hotel room of the primary case to doorway/corridor **for 1 secondary case** | Insufficient air replacement<br>Recirculating air flow |
| Gunther et al, 2020<sup>22</sup><br><u>Quality rating:</u><br>high | Meat processing plant, Germany<br>April-June 2020 | 31 confirmed cases | Genomic sequencing<br>On-site visit (work condition and ventilation system) and information provided by employer on housing, commuting and workplaces of employees | Close contact: possible for some cases<br>Fomite transmission: possible for some cases<br>Transmission from outside of event: unlikely | **Probable** airborne transmission possible for some cases on the production line, up to 12m from the primary case | Insufficient air replacement<br>Recirculating air flow from air circulation system |
| Hamner et al, 2020<sup>23</sup><br><u>Quality rating:</u><br>Low | Singing event, United States<br>10 March 2020 | 52 cases (32 confirmed and 20 probable) | No genomic sequencing<br>Telephone interviews | Close contact: possible for some cases<br>Fomite transmission: unlikely<br>Transmission from outside of event: possible for some cases | **Possible** airborne transmission >2m for some cases, due to high SAR (2.5h) | Singing<br>Insufficient air replacement (unclear)<br>Recirculating air flow from heating system (unclear) |
| Hwang et al, 2020<sup>24</sup><br><u>Quality rating:</u><br>Low | Apartment block, South Korea<br>August 2020 | 10 confirmed cases from 7 households | No genomic sequencing<br>Epidemiological data, surface sampling, building assessment | Close contact: possible for some cases<br>Fomite transmission: unlikely<br>Transmission from outside of event: possible | **Possible** airborne transmission through ventilation shaft across 11 floors | Recirculating air flow through vertical air duct/floor drain<br>Insufficient air replacement (unclear) |
| Jiang et al, 2021<sup>25</sup><br><u>Quality rating:</u> low | Department store, China<br>January 2020 | 24 confirmed cases | No genomic sequencing<br>Epidemiological data, surveillance video, assessment of ventilation conditions | Close contact: possible for some cases<br>Fomite transmission: possible<br>Transmission from outside of event: possible | **Unclear** airborne transmission across different sections of the store | Insufficient air replacement |
| Katellaris et al, 2021<sup>26</sup><br><u>Quality rating:</u> High | Singing event, Australia<br>July 2020 | 13 confirmed cases | Genomic sequencing<br>Interviews with cases, video recording, on-site visit (ventilation system) | Close contact: unlikely (except for 5 cases from same household)<br>Fomite transmission: unlikely<br>Transmission from outside of event: unlikely | **Probable** airborne transmission with secondary cases seated 1-15m from the primary case (1h) | Insufficient air replacement<br>Singing |
| Kwon et al, 2020<sup>27</sup><br><u>Quality rating:</u> high | Restaurant, South Korea<br>June 2020 | 3 confirmed cases | Genomic sequencing<br>Contact tracing, interviews, credit card records, video recording, mobile phone location data, on-site visits, air flow measurement, environmental sampling | Close contact: unlikely<br>Fomite transmission: unlikely<br>Transmission from outside of event: unlikely | **Probable** airborne transmission between cases seated 4.8m (21min) and 6.5m (5min) from primary case | Insufficient air replacement<br>Recirculating air flow by air circulation units |
| Li et al, 2021<sup>28</sup><br><u>Quality rating:</u> medium | Restaurant, China<br>January 2020 | 10 confirmed cases from 3 tables | No genomic sequencing<br>Epidemiological data, video recording, on-site visit, design of air conditioning and ventilation system, simulation and experiments to assess airflow and ventilation rates | Close contact: possible for cases within same household<br>Fomite transmission: possible<br>Transmission from outside of event: possible | **Possible** airborne transmission between primary case and at least 2 secondary cases (1/each table); up to 1.4m (53min) and 4.6m (75min) from primary case | Insufficient air replacement<br>Recirculating air flow by air circulation units |
| Lin et al, 2021<sup>29</sup><br><u>Quality rating:</u> medium | Apartment block, China<br>January 2020 | 9 confirmed cases from 3 households | Genomic sequencing<br>Interviews with cases, CCTV of the elevator, tracer gas and wind speed experiments | Close contact: likely for transmission within households<br>Fomite transmission: possible for transmission within households<br>Transmission from outside of event: unlikely | **Probable** airborne transmission between cases in 1 apartment to 2 different apartments (up to 10 floors from apartment of primary case) | Insufficient air replacement<br>Recirculating air flow through drainage and exhaust system |
| Luo et al, 2020<sup>30</sup><br><u>Quality rating:</u><br>medium | Buses, China<br>January 2020 | 9 confirmed cases | No genomic sequencing<br>Epidemiological survey, information on loading and unloading stops of all passengers | Close contact: unlikely<br>Fomite transmission: possible<br>Transmission from outside of event: possible | **Possible** long-distance (~2m) airborne transmission between some of the cases (1h to 2.5h), but **unclear** for cases 4.5m away from primary case | Insufficient air replacement<br>Recirculating air flow due to exhaust system |
| Shah et al, 2021<sup>31</sup><br><u>Quality rating:</u><br>Medium | 5 singing events, Netherlands<br>September-October 2020 | 50 cases (48 confirmed and 2 probable) | Genomic sequencing for 7 cases<br>Phone and email correspondence, questionnaires, epidemiological data, aerosol transmission model | Close contact: possible for some case<br>Fomite transmission: unlikely (except for 1 event)<br>Transmission from outside of event: possible for some cases, but unlikely in other events | **Possible** airborne transmission >1.5m for some of the cases (1h to 2.5h) | Singing<br>Recirculating air flow unclear (unclear)<br>Air replacement: unclear (unclear) |
| Shen et al, 2020<sup>32</sup><br><u>Quality rating:</u><br>Medium | Buses, China<br>January 2020 | 24 confirmed cases | No genomic sequencing<br>Questionnaires and interviews, contact tracing data, bus design and ventilation system | Close contact: possible for some cases<br>Fomite transmission: possible for some cases<br>Transmission from outside of event: unlikely | **Possible** airborne transmission >2m from primary case (50min) | Insufficient air replacement<br>Recirculating air flow from central heating system set to heating and recirculation mode |

\* The scale 'probable', 'possible', 'unlikely' and 'unclear' was used to assess the likelihood of each mode of transmission, based on study results and our critical appraisal of the study.
† Our assessment of the likelihood of airborne transmission of SARS-CoV-2 over distances greater than 2m is based on likelihood of occurring in some, but not necessarily all, transmission events.
‡ Exposure distance and time are stated where known. If not stated, exposure distance and/or time are not clear or not specified.

### Quality assessment

Detail of the quality rating is provided in Table 2, including the quality ratings: three studies rated as high quality,^22, 26, 27^ six as medium,^21, 28-32^ and four as low.^20, 23-25^

**Table 2.**
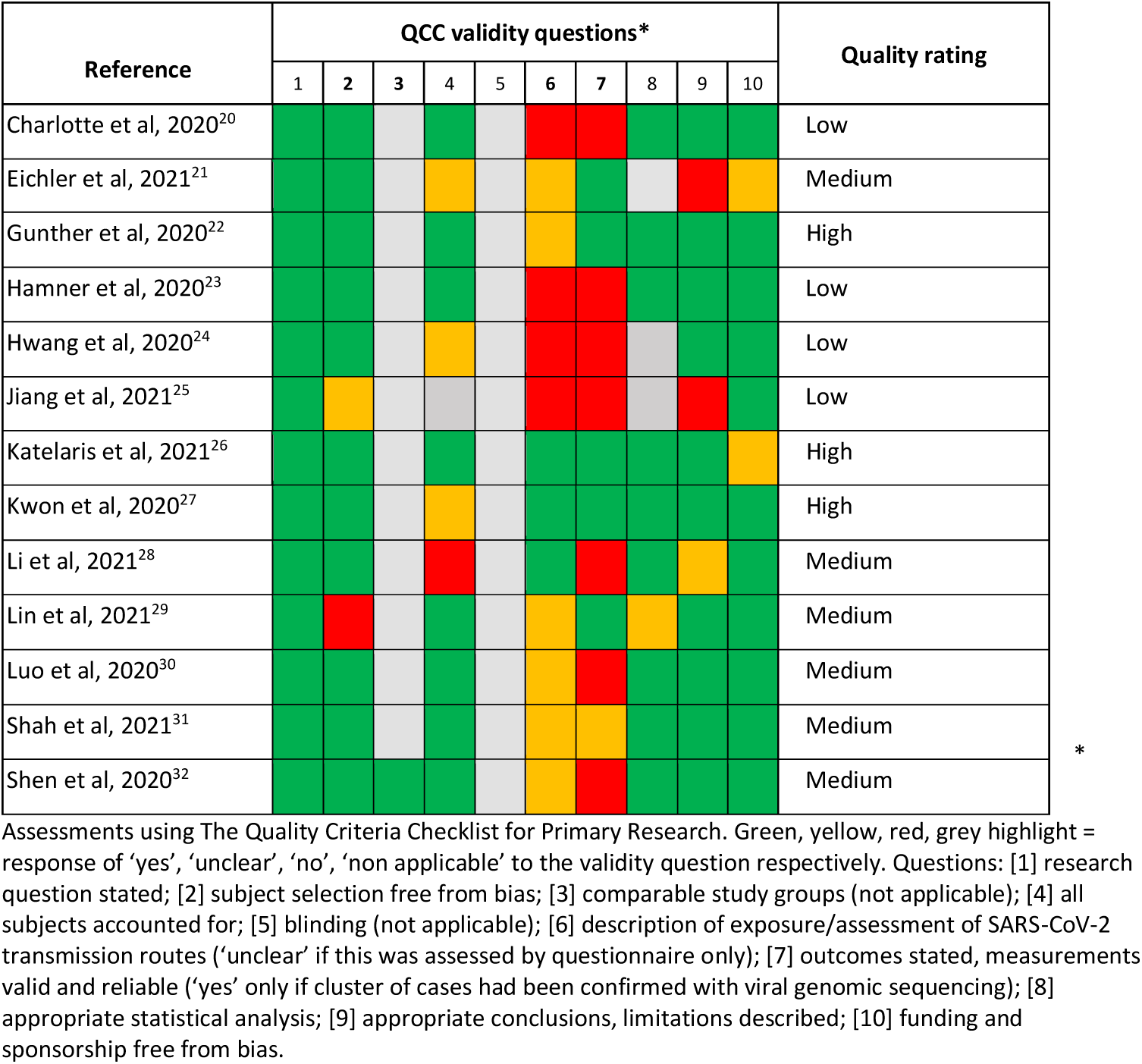
Quality assessment.

### Transmission within restaurants

Two studies reporting on two separate outbreaks within restaurants were identified; one in Jeoniu, South Korea,^27^ and one in Guangzhou, China.^28, 34^ After extensive epidemiological and environmental investigations, both studies suggested the most plausible routes of transmission of SARS-CoV-2 to be airborne at distances greater than two metres. They also concluded that air flow from air circulation units inside the restaurants combined with lack of air replacement contributed to these transmission events.

The epidemiological investigation by Kwon et al^27^ (rated as high quality) identified a transmission event at a restaurant on 12 June 2020, where three confirmed cases of COVID-19 had been visiting at the same time and were found to share the same SARS-CoV-2 genomic type using genome sequencing. The investigation, which included routine video surveillance, interviews and contact tracing, revealed that the two secondary cases had been within the restaurant at the same time as the primary case (who was presymptomatic at the time); one sat 6.5 metres away from the primary case for five minutes and another sat 4.8 metres away for 21 minutes. Extensive environmental investigation, including measurement of air flow velocity and direction, suggested that an air conditioning unit located on the ceiling had directed air from the primary case diagonally across to the two secondary cases. Neither the two staff members nor nine other visitors to the restaurant tested positive for COVID-19, including some who were closer to the primary case and for a longer period but not in the air flow path, or were facing away from the primary case. In addition, no outside ventilation system or windows in the restaurant was reported. The study authors ruled out close contact and fomite transmission based on video surveillance analysis.

An outbreak in a restaurant in China, first described by Lu et al,^34^ had further epidemiological and environmental investigations carried out and published by Li et al^28^ (rated as medium quality). The transmission event, that happened on Chinese New Year’s Eve (24 January 2020), is believed to have occurred between a primary case from Wuhan (symptom onset later that day) who sat at table A, and 9 potential secondary and tertiary cases sat at three tables (tables A, B and C). None of the other 79 visitors and eight staff tested positive or developed symptoms of COVID-19 within 14 days. The epidemiological investigation, which included travel and exposure history, suggested that at least one member of table B and one member of table C were likely to have been infected at the restaurant (the other cases could have been infected outside the restaurant as each table was constituted of family members). Results of the computational fluid dynamic simulation and of the environmental investigation, which included analysis of the air conditioning system and tracer gas measurement, found that there was a low exchange rate within the restaurant and that the three tables were located within a relatively isolated air recirculation zone at the back of the restaurant that may have promoted long-distance airborne transmission (between 1.4 and 4.6 metres). In addition, video surveillance analysis suggested that the risk of close contact and fomite transmission was low between the three tables, this was confirmed by a recent re-analysis of the video images, published after the search for this rapid review was conducted.^35^ Fomite transmission through toilet use is unlikely but not impossible. However, as no genomic sequencing was performed to confirm a connection between the cases, it is possible that transmission occurred outside of this event entirely.

### Transmission between apartments in a block

Two separate epidemiological investigations of residential clusters of COVID-19 cases within two apartment blocks were identified; one in Guangzhou, China^29^ and one in Seoul, South Korea.^24^ Both studies found a distribution of COVID-19 cases suggesting that airborne transmission of SARS-CoV-2 between apartments through vertical air ducts or floor drains was the most plausible route of transmission for these clusters of cases.

The study by Lin et al^29^ (rated as medium quality) investigated an outbreak of nine positive cases of COVID-19 in three apartments located within a 29-storey apartment block in China. The apartment block consisted of three side-by-side units (A, B and C), with one shared entrance and shared lifts in the centre of the block. The three apartments in which the cases lived were all located in unit B. Two cases from flat 15B tested positive for COVID-19 on 27 January 2020, followed by three other family members on 29 January (four of them had returned from Wuhan on 24 January 2020). The four other cases (flats 25B and 27B) tested positive between 1 and 13 February 2020 and the link between the nine cases was confirmed by genetic analysis. The apartments within the same unit shared one drain pipe, one sewer pipe and one exhaust pipe connected to each other via a ventilation pipe to the roof, although the connector in unit B was different from those in units A and C. A tracer-gas experiment showed that this change of connector may have resulted in a reduced ventilation efficiency which, combined with a lack of open windows due to cold weather, is thought to have resulted in a reduction in air replacement with fresh air. The investigators also conducted a wind speed experiment that showed that flushing a toilet caused strong airflow through the pipes and into apartments. Close contact and fomite transmissions were deemed unlikely (except for cases within same household) as COVID-19-positive residents reported no close contact with other cases, the elevator was disinfected immediately after diagnosis of the primary case and video surveillance between 25 and 27 January 2020 showed no close contact within elevators. However, only symptomatic cases were tested, therefore the potential for some transmission events to have occurred via asymptomatic cases was not considered.

A similar outbreak in a block of flats in South Korea in August 2020 was reported by Hwang et al^24^ (rated as low quality). The 437 residents from one apartment block (267 apartments) were all tested, resulting in 10 COVID-19 positive cases from seven households, spanning over 10 floors and located around two ventilation lines (eight cases within one line, two within another line). The study authors hypothesised that airborne transmission via these vertical lines of ventilation had played a role in this outbreak, although no experiments had been conducted to confirm this hypothesis. Close contact or fomite transmission in the elevator as the main transmission route was deemed improbable as this would likely have resulted in a random scattering of positive cases throughout the apartment block, rather than all being within two vertical lines. Transmission in other communal areas was also deemed unlikely as the cases reported wearing masks within these areas and no close contact between cases was reported by participants. Genomic sequencing was not performed so it is possible that transmission occurred outside of the apartment block, although again this would have likely resulted in a random distribution of cases. It is therefore possible that some of these transmission events happened via long-distance airborne transmission through the ventilation lines, particularly where eight cases from different households were located within the same vertical line of apartments.

### Transmission within buses or coaches

Two separate outbreaks in passengers on buses in China were identified, one on a bus to and from a worship event^32^ and one on a long-distance journey via public transportation coach and mini-bus.^30^ The results of both studies suggest that long-distance airborne transmission may have happened during these bus journeys in which a lack of air replacement was reported.

Following a COVID-19 outbreak among lay Buddhists who had travelled by bus to a temple in Zhejiang, China on 19 January 2020, a retrospective epidemiological investigation with a nested case-control study^32^ was carried out (rated as medium quality). A total of 31 cases were identified out of the 300 participants, of whom 128 had travelled to the event in two different buses from the same district of the city. The primary case was identified as the first to have developed symptoms and the only participant who had been in contact with individuals with travel history to Wuhan. No genomic sequencing was performed so the presence of another primary case cannot be ruled out, although this was deemed unlikely as at the time of the event most cases were thought to be contained within Wuhan. Of the 30 secondary cases, seven were thought to have been infected by close contact transmission during the religious event. The other 23 cases had travelled in the same bus as the primary case and were thought to have been mainly infected during the bus ride. Indeed, all participants had mixed during the 150-minute religious event which was mainly held outside, so transmission within the event should have resulted in a random distribution of cases rather than being largely limited to those who had travelled in the same bus as the primary case. Those travelling on the bus with the primary case were 11 times more likely to have developed COVID-19 compared to all other participants (RR 11.4; 95%CI 5.1 to 25.4; p<0.01) and 42 times more likely compared to those traveling in the other bus (RR 42.2; 95%CI 2.6 to 679.3; p<0.01). Passengers were reported as having remained seated during the bus ride and to have kept the same seats for the return journey (50 minutes each way). Cases were scattered throughout the bus and there was no significant association with being seated less than two metres from the primary case, suggesting that long-distance airborne transmission is likely to have occurred during the bus journey. The air conditioning system of the bus was on heating and recirculating mode, which may have contributed to airborne transmission through lack of air replacement. Fomite transmission, such as through a bus pole, cannot be ruled out but it is unlikely that it would have accounted for all 23 cases. The main limitations of this study, in addition to lack of genomic sequencing, are that the exposure assessment was mainly based on questionnaires and interviews with participants and that it is unclear whether all participants were tested or only those who had developed symptoms and their contacts.

Another epidemiological investigation^30^ (rated as medium quality) in China revealed transmission from one primary case to nine secondary cases that had occurred on public transport on 22 January 2020. On the day of symptom onset, the primary case had travelled without wearing a mask on a coach for 2.5 hours with 48 other individuals and then on a minibus for one hour with 12 other individuals. The secondary attack rate (SAR) was 15% (95% CI 6% to 24%) and most secondary cases were seated over two metres away (up to 4.5m both in the coach and minibus) from the primary case. Whilst fomite transmission and close contact transmission may have occurred, the investigation suggested potential for airborne transmission, in part due to the lack of air replacement (windows were closed in both buses) and increased airflow from the ventilation systems. Transmission from outside this event was deemed unlikely by the authors as there were very few COVID-19 cases in the province at the time, and none of the confirmed cases had travelled to Wuhan or been in contact with another COVID-19 case in the two weeks prior to symptom onset. However, as genomic sequencing was not performed, it is possible that some of the cases had been infected elsewhere or that there was more than one primary case. In particular, two secondary cases were potentially infectious during the coach journey (one developed symptoms and the other tested positive the day after) and could have infected other passengers, reducing the distance between primary and secondary cases. A third case, who was seated 4.5 meters away from the primary case in the minibus, could have been infected somewhere else as they developed symptoms two days after the event. This reduced the confidence that airborne transmission up to 4.5 meters may have happened in this outbreak.

### Transmission within food processing factories

An epidemiological investigation^22^ (rated as high quality) was conducted in a large meat processing complex in Germany where a COVID-19 outbreak spread throughout May and June 2020, resulting in more than 1,400 cases. The results of the retrospective investigation suggested that the outbreak started in a processing plant where 31 out of the 140 workers on the same shift tested positive for COVID-19. A genomic analysis was performed, confirming that the cases involved in this first outbreak shared the same viral genome signature and that one asymptomatic case was likely to have been the common source of infection. As most of the employees worked at a fixed position in a conveyor-belt processing line, the association between infection rate and distance to the primary case was calculated. The results show that the number of positive cases at distances between 5 and 12 metres from the primary case was significantly higher than expected for a random spatial distribution of positive cases, reaching a maximum significance level at 8 metres. The authors hypothesised that factors such as increased respiratory rates (due to physically demanding work), lack of air replacement and continuous recirculation of cooled unfiltered air might have promoted long-distance airborne transmission but this was not investigated. As some employees lived in shared accommodation and shared cars to travel to work, possible transmission routes outside the processing plant were investigated by assessing the infection rates across the shared apartments (n=11), bedrooms (n=16) and cars (n=6) and comparing them to expected rates for a random distribution of cases. The results were significant only for one shared apartment and corresponding shared car and one shared bedroom, suggesting that most transmission events occurred in the processing plant. Close contact or fomite transmission in other areas of the processing plant, such as canteens or toilets, is possible although the spatial distribution of the cases suggest that transmission is likely to have occurred on the processing line.

### Transmission in a quarantine hotel

An outbreak investigation^21^ in New Zealand (rated as medium quality) was launched in September 2020 to investigate a chain of transmission that involved nine cases, including on an international flight (26 August 2020) and a quarantine hotel (27 August-11 September 2020). By combining epidemiological investigation techniques and viral genomic sequence analyses, the primary case was identified and the genomic link between the nine cases confirmed. Close contact transmission was determined to be the most likely route for most transmission, except between cases C and D. During the 14-day quarantine, mandatory RT-PCR tests were performed on days 3 and 12. Case C tested negative on day 3 and positive on day 12 (symptom onset on day 10) and it was postulated that they had been infected during the international flight during which they were seated within two rows of the primary case. Case D, and their child case E (likely to have been infected by close contact by case D), were negative at days 3 and 12 and were identified as cases only 10 days after the end of the quarantine, on 21 September 2020, when they were tested as part of this investigation. The genomic analysis and timing of events suggest that case D was infected by case C at the quarantine hotel where they were staying in adjacent rooms. Close contact transmission between cases C and D was rejected as video surveillance showed that cases D and E were never outside their room at the same time as case C. However, video surveillance showed that when testing was performed on day 12, there was a 50 second window between case C’s door being closed and case D’s door being opened. The hotel corridor outside of the rooms was enclosed and unventilated, so lacking outside air replacement, and an assessment of the ventilation system showed the hotel room ventilation system resulted in a net positive pressure in the rooms compared to the corridor. Therefore, air containing potentially infected respiratory particles could have moved from the hotel room of the primary case into the corridor. There was a communal bin used by both cases C and D but as video surveillance showed case D had touched the bin over 20 hours after case C, the study authors considered that fomite transmission was unlikely. However, it is possible that SARS-CoV-2 can survive on non-porous surfaces for several days,^36^ so fomite transmission cannot be completely rejected. Similarly, not enough information was reported in the study to be able to fully reject close contact transmission between case D and other cases of the cluster.

### Transmission within a department store

An epidemiological investigation^25^ (rated as low quality) of an outbreak in Tianjin, China, identified a cluster of 24 cases of COVID-19 linked to a department store (6 staff and 18 customers) where transmission is believed to have happened between the 20 and 25 January 2020. Suspected routes of transmission were investigated by combining video surveillance, interviews with cases and contacts, and assessment of the ventilation conditions in the store. The primary case was thought to be a staff member with symptom onset on the 21 January 2020 who infected two other staff members who developed symptoms on 22 and 25 January 2020. There was no relationship between these three individuals and analysis of video surveillance revealed no close contact between them, including during lunch or use of toilets. Each of these three staff worked in a different department of the store, separated by a 1.5 metre corridor, and airborne transmission was considered as the most likely transmission routes by the authors. For the remaining cases, the plausible transmission routes were reported but without specifying the results of the investigation: for 10 of them, airborne transmission was deemed to be the most likely route, although distance was not specified; for five cases, droplet or fomite transmission was deemed most likely; for the remaining six cases, transmission routes could not be determined. The investigation suggested that lack of air replacement (doors were closed) and high population density in the store might have promoted airborne transmission. However, as genomic sequencing of SARS-CoV-2 was not performed, transmission outside this event cannot be ruled out. In addition, as one of the staff members experienced symptom onset the day after the suspected primary case, it is possible that several index cases were present and/or that transmission had happened earlier than what was postulated by the investigation team. Based on this investigation, it is unclear whether long-distance airborne transmission had occurred within the store.

### Transmission during singing events

Four epidemiological investigations reporting on outbreaks linked to singing events in Australia,^26^ the Netherlands,^31^ France^20^ and the USA^23^ were identified. The results from the four studies suggest that long-distance airborne transmission is likely to have happened for at least some of the transmission events, and that singing may have increased the amount of respiratory particles generated by the primary cases.

An epidemiological investigation^26^ (rated as high quality) reported on an outbreak in Sydney, Australia, linked to a series of four church services held between 15 and 17 July 2020 (one service each on the 15 and 16 July, two services on the 17 July; all in the same church). The probable primary case, a choir member, had sung at each of these one-hour services and reported symptom onset between 16 and 17 July 2020. Following the notification of two secondary cases on the 20 July, the local public health team classified all attendees at the four services as close contacts (n=508) and an on-site testing clinic was established, resulting in an 85% testing uptake. In total, 12 secondary cases were identified (2.4% SAR across the four services), all of whom had attended services on the 15 and/or 16 July 2020. No cases who had attended only on the 17 July were identified. Viral genomic sequencing of the primary case and 10 secondary cases showed that they all formed a single genomic cluster, suggesting that transmission had occurred during the church services. Seating arrangement during the services was determined by video surveillance and confirmed by interviews with the cases, showing a spatial clustering: all secondary cases had sat in the same section of the church, between 1 and 15 metres from the primary case, who was located in a choir loft 3.5 metres above the congregation, facing away from the section where the secondary cases were seated. Apart from five of the secondary cases who were from the same household, close contact or fomite transmission was deemed unlikely based on the video surveillance and reports from the primary case that they had not mixed with attendees or touched objects. The investigation revealed that windows and doors had been kept closed during the events and that the ventilation system was off, resulting in a lack of air replacement. In addition, the authors hypothesised that singing may have increased the amount of aerosols generated by the primary case. However, it is unclear why no secondary cases were identified within participants who attended only on 17 July 2020. Possible reasons hypothesised by the authors were that cases from the 17 July were not detected, that the air flow on that day was different, or that the primary case had passed their peak of infectiousness.

The second epidemiological investigation^31^ (rated as medium quality; preprint), mainly based on interviews and questionnaires, reported on five singing events held between September and October 2020 in the Netherlands. At the time, national recommendations were in place to reduce COVID-19 transmission, and whilst singing in groups was allowed, physical distancing (>1.5m) and ventilation were recommended. Each singing event had between 9 and 21 attendees (78 in total for the five events), and attack rates between 53% and 74% were observed (in total, 48 confirmed cases and 2 probable). One or more probable primary cases were identified for all events, mainly based on date of symptom onset which were between 0 and 3 days after the events. Most participants reported keeping a distance of 1.5 metres between them during the singing practice, however, distance might not have been always kept during breaks and before or after the events. In addition, some members reported having travelled together to the events by car or bike and six of the cases shared households. Therefore, close contact transmission cannot be ruled out and the authors considered it possible for some of the secondary cases in three of the five events. Fomite transmission was deemed unlikely in most cases, except for one event which involved the use of a coffee machine with a push button during the break. However, due to the high SARs, fomite and close contact transmission are unlikely to have accounted for all secondary cases. Based on a risk assessment model for exposure to SARS-CoV-2 particles published elsewhere,^37^ the authors suggested that airborne transmission due to singing might have resulted in the high attack rate observed in these outbreaks if a supershedder was present (a primary case excreting elevated concentration of virus ≥ 10^10^ virus/mL of mucus). In addition, even though ventilation through opening doors or windows was reported for all events, air exchange rates were likely to have been low in at least three of the five events. However, genomic sequencing was performed only for a minority of the cases (7 out of 50 cases), so transmission outside the events cannot be ruled out and it is possible that several index cases were present at each of these events.

The two other outbreaks occurred in March 2020, that is, during the early stage of the pandemic when no mitigation measures were in place. One of them happened in France during a choral rehearsal^20^ (study rated as low quality). Of the 27 members, 19 cases were identified in the 10 days following the event, of which seven were confirmed and 12 were probable (70% attack rate). None of the attendees reported symptoms on the day of the rehearsal and several potential primary cases were identified: one member who developed symptoms the day after the event, and two who had been in contact with COVID-19 cases in the seven days before the event. The choir practiced for 2 hours in a narrow, indoor, non-ventilated space where participants seated further apart than usual but with less than 1.8 metres between each seat. There was reported to be no close contact and minimal socialising during the event however this was mainly based on an interview conducted with the choir president two months after the event. In addition, no genomic sequencing was performed so it is possible that at least some transmission events could have occurred outside of the rehearsal as levels of community transmission were likely to have been high at the time. However, the high attack rate suggests that some airborne transmission at distances greater than two metres may have occurred.

An outbreak following a 2.5 hours choral rehearsal on 10 March 2020 in Washington (USA) was initially reported by Hamner et al^23^ (rated as low quality), and further discussed by Miller et al.^33^ One probable primary case, who had become symptomatic three days before the event, was identified and 52 of the 61 participants were identified as secondary cases (32 confirmed and 20 suspected cases; 53 and 87% SAR, respectively). The estimated space between each participant was approximately 0.75 metres lateral and 1.4 metres longitudinal (some empty seats but no specific patterns), except during a 50-minute practice when half of the group sat next to one another. Close contact transmission between some participants during this session can therefore not be ruled out. Close contact transmission during the rest of the practice and fomite transmission were both deemed unlikely by the study authors based on interviews with choir members. In both publications it was reported that singing may have increased transmission risk, which is consistent with a modelling aerosol infection risk reported by Miller et al.^33^ Doors were closed during the event but it is not known whether the ventilation and heated systems were operating. Ten of the secondary cases developed symptoms in the two days following the rehearsal (3 on 11 March and 7 on 12 March); as no viral genomic sequencing was performed, it is possible that that some of the secondary cases were infected elsewhere or that several primary cases were present. However, the high SAR suggests that long-distance airborne transmission may have occurred for at least some of the cases as not all secondary cases would have been seated near to a primary case.

### Summary and critical analysis of the results

The assessment of the likelihood of long-distance airborne transmission of SARS-CoV-2 is summarised for each study in Table 1.

Seven of the outbreaks identified^20, 23, 25, 28-30, 32^ occurred in the early stage of the pandemic (January-March 2020) when knowledge on COVID-19 was limited, especially in relation to the incubation period of SARS-CoV-2 and asymptomatic or presymptomatic transmission. As a result, most of these studies only conducted symptomatic testing and considered potential secondary cases participants with symptom onset soon after the potential exposure event, including the day after. In addition, for the studies conducted in January 2020 in China and in March 2020 in Europe or the US, it is possible that community transmission was higher than was thought at the time.

Therefore, in an outbreak such as the one reported by Luo et al^30^ where no genomic sequencing was conducted and three of the nine secondary cases developed symptoms or were tested positive one or two days after exposure, it is plausible that more than one primary case was present and that transmission occurred through means other than long-distance airborne transmission. Similarly, the evidence for long-distance airborne transmission for the outbreak in the department store was judged as unclear.^25^ In two of the studies reporting on singing events,^20, 23^ genomic sequencing and asymptomatic testing were not carried out and some of the secondary cases developed symptoms in the days following exposure. However, due to the high attack rates reported for these outbreaks, it is possible that long-distance airborne transmission had happened for at least some of the transmission events. Long-distance airborne transmission was also considered possible for two other early studies due to the detailed epidemiological investigation.^28, 32^ The study by Lin et al^29^ is the only early outbreak investigation that conducted genomic sequencing and where long-distance airborne transmission was deemed as possible.

Amongst the other studies, three^22, 26, 27^ provided convincing evidence for long-distance airborne transmission due to the detailed epidemiological investigation combined with genomic sequencing. Eichler et al^21^ also conducted genomic sequencing but their reporting of the epidemiological investigation was not sufficiently exhaustive to exclude other transmission routes (close contact or fomite) for the only secondary case that could have been infected by long-distance airborne transmission. Finally, the investigations by Shah et al^31^ and Hwang et al^24^ suggested that long-distance airborne transmission was possible for at least some of the transmission events.

### Grading of the evidence

Grading of the evidence is presented in Table 3 for each of the primary outcomes: COVID-19 infection via airborne transmission at a distance greater than two metres, insufficient air replacement (modifying factor), recirculating air flow (modifying factor) and singing (modifying factor).

**Table 3.**
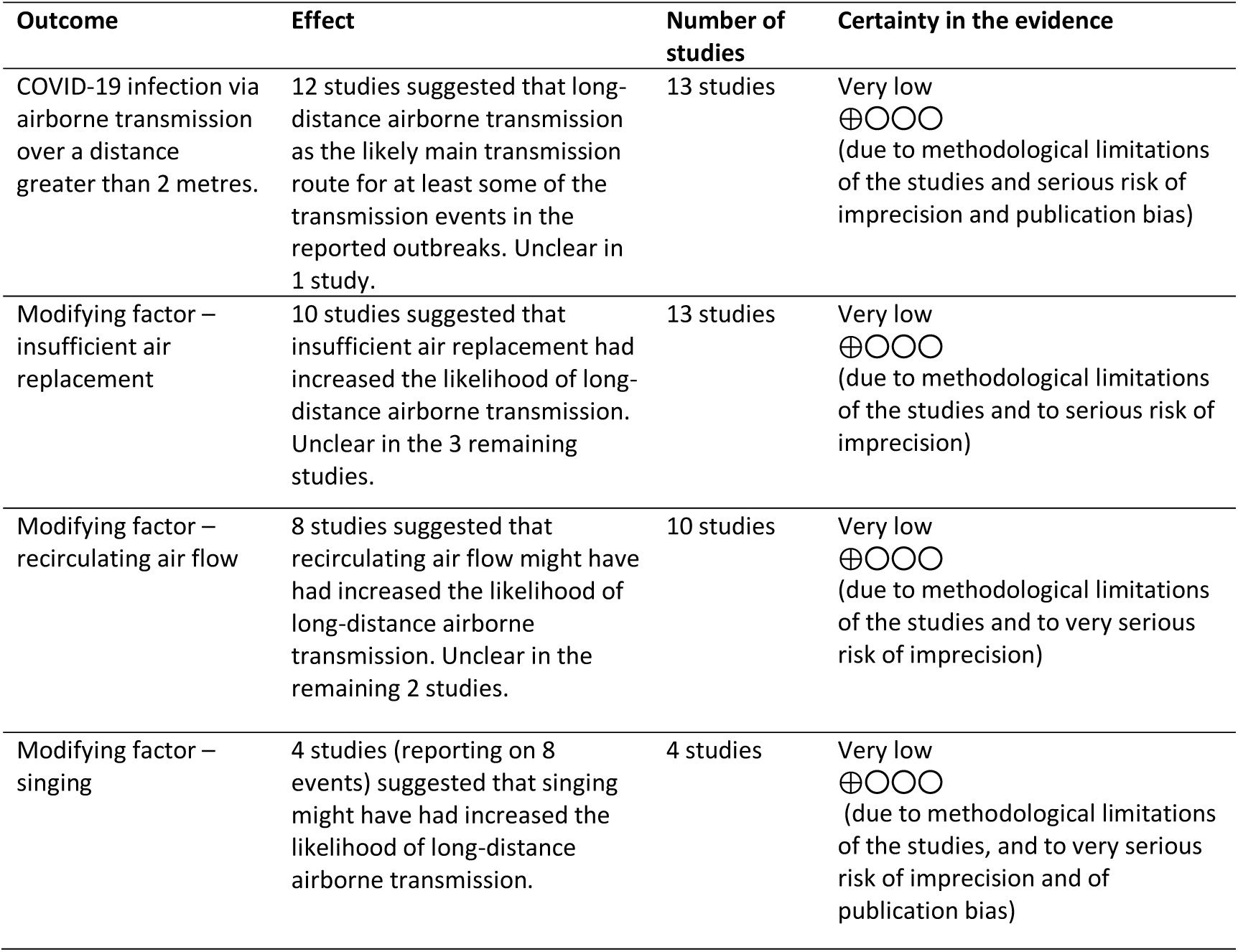
GRADE summary of findings.

For all four outcomes, the evidence was judged as having serious methodological limitations due to study design, and to have serious or very serious risk of imprecision as most studies had some risk of bias in exposure and/or outcome assessment. However, the risks of inconsistency and indirectness were judged as not serious as, despite the risk of imprecision, the results were consistent across studies conducted in a range of settings, with different populations, and provide evidence of direct relevance to the public health question of interest. The risk of publication bias was judged to be serious for the outcome COVID-19 infection via airborne transmission at a distance greater than two metres and very serious for singing, but not serious for insufficient air replacement and recirculating air flow. As a result, the certainty of evidence was judged as very low for all outcomes.

Due to high heterogeneity between studies, the additional outcomes of time spent in the transmission setting and distance over which airborne transmission was thought to have occurred could not be summarised or GRADEd. Exposure timings ranged from 5 minutes to 2.5 hours, and distances were up to 15 metres.

## DISCUSSION

Evidence from epidemiological investigations suggests that airborne transmission of SARS-CoV-2 from an infectious individual to others who were located more than two metres away can occur in different indoor community settings. However, the evidence was deemed to be of very low certainty based on 13 epidemiological investigations. The relatively small number of studies identified may suggest that outbreaks due to long-distance airborne transmission are relatively rare, although it could also be due to difficulties in identifying such events or to under-reporting, especially in countries without rigorous systems for investigating transmission routes from outbreaks or sufficient contact tracing. Indeed, the thoroughness of epidemiological investigations varied considerably between countries. In South Korea for example, investigations involved personal interviews, credit card records, analysis of video surveillance, mobile phone location data and viral genomic sequencing while in countries such as France or United States they mainly relied on questionnaires.

Interestingly, the results of this review show that when long-distance transmission occurred, one or more factors were thought to have contributed. Modifying factors such as insufficient air replacement and singing are likely to result in an increased concentration of infectious respiratory particles within the indoor space while factors such as recirculating air flow are likely to increase directional movement of air containing virus allowing viable virus to travel further. No evidence of long-distance airborne transmission occurring without one or more of these factors present was identified.

The potential for ventilation to reduce the risk of COVID-19 transmission has been acknowledged in public health guidance.^38, 39^ The results of this review confirm the importance of the role of ventilation to mitigate the risk of long-distance aerosol transmission, although caution should be taken with using recirculating air units.

In addition, a total of eight singing events (from four studies) were identified in which singing may have had increased the likelihood of long-distance airborne transmission. These results are in line with experimental and modelling studies that have reported on singing and aerosol generation, suggesting that more virus is emitted when singing compared to speaking or breathing, and may also be associated with duration and loudness of the vocalisation.^40^ More generally, the quantity of aerosol particles emitted increase with voice loudness,^41^ which was thought to have contributed to long-distance aerosol transmission in the food processing factory.^42^

In 9 out of 13 studies identified in this review, ^20, 22, 25-28, 30-32^ primary cases were asymptomatic, presymptomatic or near the time of symptom onset when transmission occurred. This finding is consistent with wider evidence that asymptomatic or presymptomatic COVID-19 cases can contribute to the community spread of COVID-19,^43^ although the exact extent is unclear.^44, 45^ The results from this review add to the evidence that asymptomatic and presymptomatic cases can contribute to long-distance airborne transmission.

Apart from one study with a nested case-control analysis, all included studies were descriptive observational studies and are therefore at high risk of bias by design, including risk of recall bias and lack of control group. In addition, outbreak investigations often have small sample sizes which are not necessarily representative and so may lack generalisability. Nonetheless, these findings are an important addition to the wider body of evidence that supports the biological plausibility of airborne transmission as a potentially significant route of transmission in certain scenarios. The wider evidence includes experimental studies that have shown that SARS-CoV-2 can remain viable in artificially generated aerosols for up to 16 hours, and that the stability and viability depends on environmental factors such as temperature, humidity and sunlight exposure.^11, 46^ Similarly, biological monitoring studies have shown that SARS-CoV-2 RNA can be detected in exhaled breath and environmental air samples, but the evidence on viable virus is limited to a few studies that mostly detected infectious virus in air samples collected at less than two metres from the infectious individual.^11, 46^ These experimental and biological studies provide evidence that SARS-CoV-2 can be viable in aerosols and therefore support the epidemiological evidence from this rapid review, and others^46^ that suggest that airborne transmission can happen in some settings.

### Strengths and limitations

This rapid systematic review is the first to critically assess the likelihood of long-distance airborne transmission of SARS-CoV-2 using only direct real-world evidence from observational studies in indoor community settings. The risk associated with close contact transmission is now relatively well-known, at least in indoor settings, but there remains a need to further understand long-distance transmission to support public health decision making, especially in relation to the implementation of effective and proportionate mitigation measures in indoor community settings.

Another strength of this review is the application of inclusion criteria that focused the critical appraisal on those studies which involved comprehensive epidemiological investigations. Some of these studies did not only include epidemiological data, but also genomic analysis, video surveillance, analysis of seating arrangements and environmental hypothesis testing. This has enabled this synthesis to provide the best evidence currently available to examine the potential for long-range airborne transmission in real-world settings.

The main limitation of selecting studies of only real-world human-to-human transmission events is that scenarios where transmission has not occurred will not be reported, and likewise where transmission events have not been detected by contact tracing systems - this could be seen as a form of publication bias. All of the evidence is from retrospective epidemiological investigations of outbreaks and therefore this review cannot make inferences on the extent to which long-distance airborne transmission occurs and the contribution it may have on community rates of transmission. Another limitation of the review is that only 10% of the results of the literature search were screened in duplicate, although with good agreement (>97%). Finally, and as with all reviews assessing COVID-19 evidence, this rapid review is limited by the fact that the evidence assessed is from an emerging field that spans over about 14 months. Although only one of the included studies was a preprint, studies conducted in the COVID-19 context are conducted at pace with the aim to provide evidence in a timely manner, which can affect the quality of the studies, both in term of design (mainly descriptive epidemiological studies) and reporting (insufficient detail provided).

### Policy implications

The results from this rapid systematic review highlight the need to ensure measures to mitigate against SARS-CoV-2 long-distance transmission in indoor settings, especially in poorly ventilated spaces. Identification of poorly ventilated public spaces should be undertaken and improvements made. Other factors such as directional air flow or singing may increase the risk for long-distance airborne transmission.

## CONCLUSION

This rapid review found evidence of long-distance (greater than two metres) airborne human-to-human transmission of SARS-CoV-2 within indoor community settings, which can occur from asymptomatic or presymptomatic cases. All transmission events identified were found to occur alongside factors believed to have contributed to this type of transmission, including lack of air replacement (absence or little ventilation with fresh air), increased recirculating air flow (mainly through air circulation systems) and singing. No studies were identified reporting the occurrence of long-distance airborne transmission without one or more of these factors present.

This highlights the importance of assessing ventilation, especially in indoor spaces where people meet others from outside of their household. Particular attention should be given to ventilation in settings with activities that may increase the number of respiratory particles, for example, singing. Where ventilation is assessed to be inadequate, improvements should be made.

We thank colleagues within UKHSA for their support into specific aspects of this review, especially Bethany Walters and Marialena Trivella. JP is supported by a British Heart Foundation Accelerator Award (AA/18/7/34219) and works in a Unit that is supported by the University of Bristol and UK Medical Research Council (MC_UU_00011/6), and was also supported by a secondment to the COVID-19 Rapid Evidence Service.

## Data Availability

Data are from published research and therefore are in the public domain. Extracted data are available in the supplementary material.

## Contributors

JP, RC, NPS and DD designed the review, with input from EOC and AB. NPS conducted the literature search. JP, DD, IT and JKSA conducted the screening and the data extraction. JP and DD conducted the critical appraisal, interpreted the findings and drafted the manuscript, with further input from RC, EOC and AB. RC supervised the study and is the study guarantor. All authors had direct access to the data, or access was provided as requested. The corresponding author attests that all listed authors meet authorship criteria and that no others meeting the criteria have been omitted.

## Ethical approval

Not required

## Data sharing

The lead author affirms that the manuscript is an honest, accurate, and transparent account of the study being reported; that no important aspects of the study have been omitted; and that any discrepancies from the study as planned (and, if relevant, registered) have been explained.

## Dissemination to participants and related patient and public communities

This paper will be shared widely within the UK Health Security Agency (UKHSA) and used to inform relevant guidance. It will also be shared with relevant stakeholders, policy makers and other public health agencies within the UK and internationally. Further dissemination will include our website https://ukhsalibrary.koha-ptfs.co.uk/covid19rapidreviews/ and via colleagues working in COVID-19 evidence synthesis.

## Provenance and peer review

Not commissioned; externally peer reviewed

