## Supplementary for "Airborne transmission of SARS-CoV-2 over distances greater than two metres: a rapid systematic review"

**Supplementary material:**

**Supplementary Figure 1. Search strategy Ovid Medline**

1. exp coronavirus/
2. exp Coronavirus Infections/
3. ((corona* or corono*) adj1 (virus* or viral* or virinae*)).ti,ab,kw.
4. (coronavirus* or coronovirus* or coronavirinae* or CoV or HCoV*).ti,ab,kw.
5. covid*.nm.
6. (2019-nCoV or 2019nCoV or nCoV2019 or nCoV-2019 or COVID-19 or COVID19 or CORVID-19 or CORVID19 or WN-CoV or WNCoV or HCoV-19 or HCoV19 or 2019 novel* or Ncov or n-cov or SARS-CoV-2 or SARSCoV-2 or SARSCoV2 or SARS-CoV2 or SARSCov19 or SARS-Cov19 or SARSCov-19 or SARS-Cov-19 or Ncovor or Ncorona* or Ncorono* or NcovWuhan* or NcovHubei* or NcovChina* or NcovChinese* or SARS2 or SARS-2 or SARScoronavirus2 or SARS-coronavirus-2 or SARScoronavirus 2 or SARS coronavirus2 or SARScoronovirus2 or SARS-coronovirus-2 or SARScoronovirus 2 or SARS coronovirus2).ti,ab,kw.
7. (respiratory* adj2 (symptom* or disease* or illness* or condition*) adj10 (Wuhan* or Hubei* or China* or Chinese* or Huanan*)).ti,ab,kw.
8. ((seafood market* or food market* or pneumonia*) adj10 (Wuhan* or Hubei* or China* or Chinese* or Huanan*)).ti,ab,kw.
9. ((outbreak* or wildlife* or pandemic* or epidemic*) adj1 (Wuhan* or Hubei or China* or Chinese* or Huanan*)).ti,ab,kw.
10. or/1-9
11. (aerosol or aerosols or aerosolized or aerosolised or airborne).ti.
12. (airborne or aerosol or aerosols or aerosolized or aerosolised or air flow* or aerodynamic* or air condition* or droplet* or cough* or sneez* or breath* or sing or singing or shout* or (air adj2 circulat*) or air recirculation or ((viral or virus) adj2 particle*)).tw,kw.
13. (transmission or distanc* or dispersal or dispersion).tw,kw.
14. 12 and 13
15. (ventilation and (transmission or distanc*)).tw,kw.
16. ((route or mode) adj2 transmission).tw,kw.
17. ((far-field or far field) and (exposure or transmission)).tw,kw.
18. Disease Transmission, Infectious/
19. 12 and 18
20. Ventilation/
21. Air Conditioning/
22. 11 or 14 or 15 or 16 or 17 or 19 or 20 or 21
23. 10 and 22
24. 23 not (exp animals/ not humans.sh.)
25. limit 24 to english language

**Supplementary Table 1. Excluded studies with reason for exclusion**

| Retrieval method | Reference | Reason for exclusion |
|---|---|---|
| Literature search | Azimi, P. et al, Mechanistic transmission modeling of COVID-19 on the Diamond Princess cruise ship demonstrates the importance of aerosol transmission | Wrong study design |
| | Bielecki, M. et al, Social Distancing Alters the Clinical Course of COVID-19 in Young Adults: A Comparative Cohort Study | Cannot ascertain exposure |
| | Bohmer, M. M. et al, Investigation of a COVID-19 outbreak in Germany resulting from a single travel-associated primary case: a case series | Cannot ascertain exposure |
| | Chau, N. V. V. et al, Superspreading Event of SARS-CoV-2 Infection at a Bar, Ho Chi Minh City, Vietnam | Cannot ascertain exposure |
| | Chen, X. et al, Clinical features and short-term outcomes of patients with COVID-19 due to different exposure history | Cannot ascertain exposure |
| | Deresinski, S., Possible Aerosol Spread of SARS-CoV-2 in an Apartment Building | Wrong study design |
| | Driessche, K.V. et al, Exposure to cough aerosols and development of pulmonary COVID-19 | Wrong exposure |
| | Ehrhardt, J. et al, Transmission of SARS-CoV-2 in children aged 0 to 19 years in childcare facilities and schools after their reopening in May 2020, Baden-Wurttemberg, Germany | Cannot ascertain exposure |
| | Evangelista, H. et al, Combining science and social engagement against Covid-19 in a Brazilian Slum | Wrong study design |
| | He, F. et al, Comparative Analysis of 95 Patients with Different Severity in the Early Outbreak of COVID-19 in Wuhan, China | Cannot ascertain exposure |
| | Ho, C., Modelling Airborne Transmission and Ventilation Impacts of a COVID-19 Outbreak in a Restaurant in Guangzhou, China | Wrong study design |
| | Kang, Y. et al, A retrospective view of pediatric cases infected with SARS-CoV-2 of a middle-sized city in mainland China | Cannot ascertain exposure |
| | Kim, J. G. et al, Air evacuation of passengers with potential SARS-CoV-2 infection under the guidelines for appropriate infection control and prevention | Wrong exposure |
| | Kolinski, J. M, et al, Superspreading events suggest aerosol transmission of SARS-CoV-2 by accumulation in enclosed spaces | Wrong study design |
| | Kristiansen, M. F. et al, Epidemiology and clinical course of first wave coronavirus disease cases, faroe islands | Wrong exposure |
| | Kwon, K. S. et al, Erratum: Correction of Text in the Article "Evidence of Long-Distance Droplet Transmission of SARS-CoV-2 by Direct Air Flow in a Restaurant in Korea" | Erratum of included study |
| | Lotta-Maria, A. H. et al, Healthcare workers high COVID-19 infection rate: the source of infections and potential for respirators and surgical masks to reduce occupational infections | Cannot ascertain exposure |
| | Pokora, R. et al, Investigation of superspreading COVID-19 outbreaks events in meat and poultry processing plants in Germany: A cross-sectional study | Cannot ascertain exposure |
| | Rankin, D. A. et al, Outbreak of COVID-19 among school auction attendees: Was it a "silent auction" or "silent transmission"? | Cannot ascertain exposure |
| | Redditt, V. et al, Outbreak of SARS-CoV-2 infection at a large refugee shelter in Toronto, April 2020: a clinical and epidemiologic descriptive analysis | Cannot ascertain exposure |

| Retrieval method | Reference | Reason for exclusion |
|---|---|---|
| | Sami S. et al, Community Transmission of SARS-CoV-2 Associated with a Local Bar Opening Event - Illinois, February 2021 | Cannot ascertain exposure |
| | Sugano, N. et al, Cluster of Severe Acute Respiratory Syndrome Coronavirus 2 Infections Linked to Music Clubs in Osaka, Japan | Cannot ascertain exposure |
| | Szablewski, C. M. et al, SARS-CoV-2 Transmission and Infection Among Attendees of an Overnight Camp - Georgia, June 2020 | Cannot ascertain exposure |
| | Wada, K. et al, Infection and transmission of COVID-19 among students and teachers in schools in Japan after the reopening in June 2020 | Cannot ascertain exposure |
| | Wilburn, J. et al, COVID-19 within a large UK prison with a high number of vulnerable adults, march to june 2020: An outbreak investigation and screening event | Cannot ascertain exposure |
| | Xie, C. et al, The evidence of indirect transmission of SARS-CoV-2 reported in Guangzhou, China | Cannot ascertain exposure |
| | Xu, P. et al, Lack of cross-transmission of SARS-CoV-2 between passenger's cabins on the Diamond Princess cruise ship | Wrong study design |
| | Yuan, Y. et al, Molecular epidemiology of SARS-CoV-2 clusters caused by asymptomatic cases in Anhui Province, China | Cannot ascertain exposure |
| | Zhang, Z. et al, Disease transmission through expiratory aerosols on an urban bus | Wrong study design |
| | Zuckerman, N. S. et al, Comprehensive Analyses of SARS-CoV-2 Transmission in a Public Health Virology Laboratory | Cannot ascertain exposure |
| Comber et al, Airborne transmission of SARS-CoV-2 via aerosols. Rev Med Virol. 2020:e2184 | Almilaji, O., Air Recirculation Role in the Spread of COVID-19 Onboard the Diamond Princess Cruise Ship during a Quarantine Period | Cannot ascertain exposure |
| | Bays, D.J. et al, Investigation of nosocomial SARS-CoV-2 transmission from two patients to health care workers identifies close contact but not airborne transmission events | Wrong setting / exposure |
| | Cai, J. et al, Indirect Virus Transmission in Cluster of COVID-19 Cases, Wenzhou, China, 2020 | Cannot ascertain exposure |
| | Zhang, R. et al, Identifying airborne transmission as the dominant route for the spread of Covid-19 | Wrong study design |
| | Cheng, V.C.C. et al, Air and environmental sampling for SARS-CoV-2 around hospitalized patients with coronavirus disease 2019 (Covid-19) | Wrong study design/settings |
| | Chia, P.Y. et al, Detection of air and surface contamination by SARS-CoV-2 in hospital rooms of infected patients | Wrong study design |
| | Faridi, S et al, A field indoor air measurement of SARS-CoV-2 in the patient rooms of the largest hospital in Iran | Wrong study design |
| | Fears, A.C. et al, Persistence of severe acute respiratory syndrome coronavirus 2 in aerosol suspensions | Wrong study design |
| | Guo, Z et al, Aerosol and Surface Distribution of Severe Acute Respiratory Syndrome Coronavirus 2 in Hospital Wards, Wuhan, China, 2020 | Wrong study design |
| | Jiang, Y. et al, Clinical Data on Hospital Environmental Hygiene Monitoring and Medical Staff Protection during the Coronavirus Disease 2019 Outbreak | Wrong study design |
| | Lei, H et al, SARS-CoV-2 environmental contamination associated with persistently infected COVID-19 patients | Wrong study design |

| Retrieval method | Reference | Reason for exclusion |
|---|---|---|
| | Liu, Y. et al, Aerodynamic analysis of SARS-CoV-2 in two Wuhan hospitals | Wrong study design |
| | Ma, J. et al, Exhaled breath is a significant source of SARS-CoV-2 emission | Wrong study design |
| | Ong, S.W.X. et al, Air, surface environmental, and personal protective equipment contamination by severe acute respiratory syndrome coronavirus 2 (SARS-CoV-2) from a symptomatic patient | Wrong study design |
| | Razzini, K. et al, SARS-CoV-2 RNA detection in the air and on surfaces in the COVID-19 ward of a hospital in Milan, Italy | Wrong study design |
| | Santarpia, J.L. et al, Aerosol and Surface Transmission Potential of SARS-CoV-2 | Wrong study design |
| | Santarpia, J.L. et al, The Infectious Nature of Patient-Generated SARS-CoV-2 Aerosol | Wrong study design |
| | Schuit, M. et al, Airborne SARS-CoV-2 is rapidly inactivated by simulated sunlight | Wrong study design |
| | Van Doremalen, N. et al, Aerosol and surface stability of SARS-CoV-2 as compared with SARS-CoV-1 | Wrong study design |
| | Wong, J.C.C. et al, Environmental Contamination of SARS-CoV-2 in a Non-Healthcare Setting Revealed by Sensitive Nested RT-PCR | Wrong study design |
| | Yamagishi T. et al, Environmental sampling for severe acute respiratory syndrome coronavirus 2 (SARS-CoV-2) during a coronavirus disease (COVID-19) outbreak aboard a commercial cruise ship | Wrong study design |
| | Yu, L. et al, Catch and kill airborne SARS-CoV-2 to control spread of COVID-19 by a heated air disinfection system | Wrong study design |
| | Zhou, L. et al, Detection of SARS-CoV-2 in Exhaled Breath from COVID-19 Patients Ready for Hospital Discharge | Wrong study design |
| | Zhou, L. et al, Investigating SARS-CoV-2 surface and air contamination in an acute healthcare setting during the peak of the COVID-19 pandemic in London | Wrong settings/exposure |

**Supplementary Table 2. Data extraction of included studies**

| Author, setting, period, objective | Participants, methods | Results relating to transmission routes | Results relating to transmission modifying factors | Study quality and key limitations |
|---|---|---|---|---|
| Charlotte et al, 2020 [1]<br><br>Setting: indoor choir rehearsal in a 45m² and 3m high room, France<br><br>Study period: 2-hour rehearsal on 12 March 2020, main interview 9 May, multiple telephone interviews up to 20 June 2020<br><br>Objective: to investigate an outbreak or superspreading event believed to have occurred at a choir practice | Participants: n=27 (25 singers, 1 conductor and 1 accompanist)<br><br>Outcome assessment<br>- COVID-19 diagnostic (confirmed if nasopharyngeal swab RT-PCR positive and/or a severe case requiring hospitalization; probable if diagnosed by general practitioners but no RT-PCR test)<br><br>Exposure assessment<br>- Questionnaire to all participants (mainly about symptom and diagnostic, response rate 100%)<br>- Telephone interviews with president and conductor of choir, discussing possible exposure, participant seating arrangement (sketch, including location of cases) and hall dimensions | - 19 COVID-19 cases identified 1-12 days following the rehearsal (7 confirmed and 12 probable); overall secondary attack rate (SAR): 70%.<br>- None of the attendees presented symptoms on 12 March. Several possible primary cases: 1 with symptom onset the day after the event and possibly others who had a close contact with a COVID-19 case in the 7 days before the event and had symptom onset 2-3 days after the event.<br>- Choristers were sat less close to each other than usual, at a distance of < 6 feet (1.8m).<br>- Close contact and fomite transmission deemed unlikely based on interview with the choir president (rehearsal in one go, with minimal socialisation, no shaking hands or food sharing; participants left the room quickly after the session and no indoor side-by-side or prolonged face-to-face contacts observed). | Insufficient air replacement<br>- The room was a narrow indoor space without ventilation.<br><br>Singing<br>- May have increased the amount of aerosols generated by the primary case(s). | Quality rating: low<br><br>Key limitations<br>- Probable cases were not confirmed with a COVID-19 test and no asymptomatic testing carried out.<br>- No genomic sequencing performed, so transmission outside the event cannot be ruled out (rehearsal occurred 5 days before the first stay-at-home order in France, so community levels may have been high).<br>- High risk of recall bias as the investigation is mainly based on interview with the president of the choir, which was conducted 2 months after the event.<br>- Singing as a modifying factor for airborne transmission was presented as a hypothesis, no data to support it. |
| Eichler et al, 2021 [2]<br><br>Setting: New Zealand quarantine hotel; part | Participants: 9 COVID-19 cases (including the primary case), of which 6 had travelled on the same international flight and | - Genomic sequencing confirmed genomic link between the 9 cases. | Insufficient air replacement<br>- Corridor enclosed and unventilated.<br><br>Recirculating air flow | Quality rating: medium<br><br>Key limitations |

| Author, setting, period, objective | Participants, methods | Results relating to transmission routes | Results relating to transmission modifying factors | Study quality and key limitations |
|---|---|---|---|---|
| of the outbreak investigated<br><br><u>Study period</u>: first case identified 18 September 2020; exposure period 26 August-11 September 2020<br><br><u>Objective</u>: to investigate the origin of infection of an international arrival who had spent 14 days in a managed isolation and quarantine (MIQ) hotel prior to developing COVID-19 | were quarantined in MIQ, and 3 household contacts.<br><br><u>Outcome assessment</u><br>- COVID-19 test (nasopharyngeal swab, RT-PCR) at days 3 and 12 for all those in MIQ<br>- Regular health monitoring<br>- Genome sequencing<br><br><u>Exposure assessment</u><br>- Epidemiological data obtained via public health authorities<br>- Video surveillance (CCTV analysis)<br>- Review of ventilation system in MIQ | - Case C, symptomatic since day 10 of quarantine, tested positive on day 12 and was relocated to an isolation section.<br>- Cases D and E, who had travelled on the same flight as case C and were quarantined in adjacent room in MIQ, tested positive 10 days after the end of MIQ stay. They had had negative tests on days 3 and 12. Timeline of events and phylogenetic trees suggest that case D was infected by case C during hotel MIQ stay (rather than during flight).<br>- CCTV evidence showed that cases C, D and E were not outside their room at the same time, but that on the day 12 testing, there was a 50 second window between the door of case C being closed and the door of cases D and E being opened. Airborne transmission during this moment was hypothesised to be the most probable mode of transmission.<br>- A communal bin was touched by cases C and D, but fomite transmission considered unlikely by study authors as CCTV showed that there was > 20 hours between they each touched it.<br>- Case E (child of case D) likely to have been infected by case D in MIQ or in household settings; in both cases close contact transmission cannot be ruled out. Similarly, for all the other transmission events of this | - The hotel room ventilation system resulted in a net positive pressure in the room compared to the corridor, meaning air and aerosol particles were likely to move from the hotel room of the primary case into the corridor. | - The transmission events spans over more than 2 weeks and includes a variety of settings. The information provided in the study is not enough to rule out other transmission routes and/or that case D had been infected by a primary case other than C.<br>- The seats of cases D and E during the flight were not specified, nor were possible interaction between cases A, B and D before A and B tested positive on day 3. |

| Author, setting, period, objective | Participants, methods | Results relating to transmission routes | Results relating to transmission modifying factors | Study quality and key limitations |
|---|---|---|---|---|
| | | outbreak (in flights or in household settings), close contact transmission cannot be ruled out. | | |
| Gunther et al, 2020 [3]<br><br>Setting: beef and pork processing complex in Rheda-Wiedenbrück, Germany<br><br>Study period: May-June 2020<br><br>Objective: to report on an outbreak that occurred at a meat processing complex | Participants: n=6289 employees, of which more than 1400 tested positive between May and June 2020. Outbreak started in one of the processing lines where 31 of the 140 employees of the same shift ('early shift') tested positive.<br><br>Outcome assessment<br>- COVID-19 test (oropharyngeal swab; RT-PCR) for all workers of the early shift<br>- Genome sequencing of 20 positive cases<br><br>Exposure assessment<br>- Information on housing, commuting and workplaces of employees provided by employer<br>- On-site visit during working hours to assess work conditions, including inspection of ventilation system | - Two staff members (asymptomatic) tested positive on 21 May, one of which considered as the primary case of the cluster (supported by genome analysis). Between 27 May and 3 June, 29 of 140 other staff members on the same shift tested positive. Genome analysis showed cases related to the same sub-branch of the virus.<br>- Production line staff have fixed workplaces with exception of supervisors.<br>- P-values for the cumulative probability of infection rates among employees working at fixed positions under the null-hypothesis that infection rates were independent of spatial distribution in function of distance were calculated, showing that the probability for spatial over-representation of cases was significant at 5 to 12m from the primary case and reaches a maximum significance level at 8m (p=2.3x10-5.); only 3 of the cases were within 2m of the primary case.<br>- Shared accommodation (11 shared flats and 16 shared bedrooms) and carpools (n=6) by some of the workers of the early shift. P-values were calculated for infection rates among employees sharing one or more unit under the null-hypothesis of a random | Insufficient air replacement<br>- Re-circulation of cooled air with low rate of exchange with fresh air (air exchange rate <1; more than 1h needed to have the air replaced with fresh air); no filter.<br><br>Recirculating air flow<br>- Eight cooling fans projected air in lateral direction. | Quality rating: high<br><br>Key limitations<br>- Close contact and fomite transmission during the 2 hours of breaks, during which staff visited the canteen, cannot be ruled out.<br>- No interviews with employees conducted, no information on whether transmission within employees outside work and housing settings (e.g. during social events) was provided.<br>- Airflow direction and speed assessment were qualitative, no experiments conducted. |

| Author, setting, period, objective | Participants, methods | Results relating to transmission routes | Results relating to transmission modifying factors | Study quality and key limitations |
|---|---|---|---|---|
| | | distribution. Positive rates were significant for only 3 of the units, which also corresponds to a positive correlation between these units' infection rates and percentage of staff working within 8 metres of primary case (average Pearson correlation coefficient r = 0.67). | | |
| Hamner et al, 2020 [4]<br>Additional evidence from Miller et al, 2020[5] which reported on the same outbreak<br>Setting: choir practice in Washington, United States<br>Study period: rehearsal on 10 March 2020, investigation 18-20 March 2020, with follow-up interviews on 7-10 April 2020<br>Objective: to investigate an outbreak that occurred at choir practice | Participants: n=61 (median age: 69 years)<br>Outcome assessment<br>- COVID-19 diagnostic (confirmed if RT-PCR positive [type of swab not specified] or suspected based on symptoms)<br>Exposure assessment<br>Telephone interviews with all choir members, focusing on rehearsal attendance, other possible exposures, symptoms and seating arrangement | - One suspected primary case (symptomatic since 7 March); estimated secondary attack rates of 53% for confirmed cases (n=32) and of 87% if including suspected cases (n=52) during the choir practice on the 10 March.<br>- Symptom onset for secondary cases: median of 3 days; range 1-12 days (3 confirmed cases on 11 March; 5 confirmed and 2 probable on 12 March; 1 probable on 22 March; all other cases had symptom onset 3-7 days after event).<br>- The choir practice lasted 2.5 hours: 2 sessions of 40-45 minutes with all attendees, a 50-minute session where attendees were split into 2 groups, and a 15-minute break.<br>- Seating arrangement: for the sessions with all attendees, some empty seats but no specific spatial patterns (estimated space between attendees: 0.75m lateral distance and 1.4m forward distance); for the session in 2 groups, one group was in a smaller room where they sat next to each other. | Insufficient air replacement<br>- External doors closed.<br>- Ventilation system and heated system (with filter >1 micron) that can both provide outdoor air intake as well as recirculating air. It is not known whether it operated continually or how much external air was supplied.<br>Recirculating air flow<br>- It is not known whether the air handling unit was producing air currents.<br>Singing<br>May have increased the amount of aerosols generated by the primary case. | Quality rating: low<br>Key limitations:<br>- Suspected cases were not confirmed with a COVID-19 test and no asymptomatic testing.<br>- Genomic sequencing not performed, transmission outside this event cannot be ruled out.<br>- Risk of recall bias as mainly based on telephone interviews.<br>- Singing as a modifying factor for airborne transmission was presented as a hypothesis, no data presented to support it. |

| Author, setting, period, objective | Participants, methods | Results relating to transmission routes | Results relating to transmission modifying factors | Study quality and key limitations |
|---|---|---|---|---|
| | | - Close contact or fomite transmission during the 15-minute break, chair stacking at the end of the practice, and use of bathroom deemed unlikely by study authors.<br>- The primary case spoke minimally with other participants. Many participants arrived shortly before the rehearsal and left as soon as it had finished.<br>- The high rate of cases suggests that some airborne infection >2m may have occurred. | | |
| Hwang et al, 2020 [6]<br><u>Setting</u>: apartment block in Seoul, South Korea<br><u>Study period</u>: investigation started on 25 August 2020 after 5 cases tested positive 23-25 August. Potential exposure date not specified<br><u>Objective</u>: to investigate an outbreak that occurred along two vertical lines in an apartment block | <u>Participants</u>: 10 COVID-19 cases (7 households); 437 residents (267 households) were tested.<br><u>Outcome assessment</u><br>- COVID-19 test (nasopharyngeal swab; RT-PCR) for all residents.<br><u>Exposure assessment</u><br>- Epidemiological data, including potential source of exposure and contact with other cases in the building (not clear how collected)<br>- Surface sampling of ventilation grills and drains<br>- Assessment of the structure of the building and air ducts | - 10 cases from 7 households, spanning over 10 floors, tested positive for COVID-19; all symptomatic but one. The suspected primary case had symptom onset on 16 August, and the other cases between 18 and 25 August.<br>- Epidemiological investigation showed that cases reported no close contact between them and that all cases reported having used masks outside their apartments.<br>- All households used the same elevators (except 2 cases) and entrance halls, but transmission via elevator (fomites or droplets) would be scattered rather than in a vertical line.<br>- All infected households located within 2 vertical lines (8 cases within 1 line, 2 within another line). Each line has a ventilation shaft which runs from the bottom to the rooftop and connects to the apartments | <u>Recirculating air flow</u><br>- Airborne transmission of SARS-CoV-2 through the vertical air duct or floor drain connecting the apartments hypothesised.<br>- There were no bathroom exhaust fans and so no physical block of the air from the ventilation shafts moving into the apartment.<br><u>Insufficient air replacement</u><br>- Cannot be assessed based on the information provided in the study. | <u>Quality rating</u>: low<br><u>Key limitations</u><br>- No genomic testing performed, transmission from cases outside the apartment block cannot be ruled out.<br>- Very limited details provided on the epidemiological investigation.<br>- Surface sampling likely to have been carried out after 25 August, that is 10 days after symptom onset of the first case.<br>- No information provided on possible follow-up: symptom onset of the cases between 16 and 25 August, testing of all residents performed around 26-27 August; cannot be ruled |

| Author, setting, period, objective | Participants, methods | Results relating to transmission routes | Results relating to transmission modifying factors | Study quality and key limitations |
|---|---|---|---|---|
| | | through the blowhole in the bathroom, which could have promoted airborne transmission between apartments.<br>- Surface samples all negative (RT-PCR). | | out that some cases would have been within incubation period at that time. |
| Jiang et al, 2021 [7]<br>Setting: Baodi department store in Tianjin, China<br>Study period: exposure period 20–25 January 2020. After 3 staff tested positive, store closed on January 26 and investigation started<br>Objective: To investigate an outbreak that occurred in a department store | Participants: 24 confirmed COVID-19 cases who worked at (6) or had visited (18) the department store<br>Outcome assessment<br>- COVID-19 test (nasopharyngeal swab; RT-PCR)<br>Exposure assessment<br>- Surveillance video of the store (20-25 January)<br>- Interviews with cases and contacts, including contact history<br>- Map of the department store and assessment of ventilation conditions (not specified how this was obtained) | - Primary case identified as a staff member, with symptom onset on 21 January 2020.<br>- Two other staff members, with symptom onset on 22 and 25 January, likely to have been infected by primary case. No relationship reported between any of the 3 staff, and no close contact behaviours (video surveillance). Most departments of the store were separated by a 1.5m wide corridor and salespersons worked in a specific area; long-distance airborne transmission likely route of transmission.<br>- Similarly, for 10 other cases (3 staff and 7 customers), airborne transmission was deemed as the most likely route of transmission.<br>- For 5 customers, droplet or contact transmission most likely.<br>- For 6 customers, transmission route could not be determined. | Insufficient air replacement<br>- Doors were closed with draft excluders to keep the store warm in the winter conditions.<br>- No air conditioning system in use. | Quality rating: low<br>Key limitations:<br>- No genomic sequencing, transmission outside the event cannot be ruled out.<br>- The 24 cases were part of a wider investigation of 131 cases. Epidemiological results traced back these 24 cases to the store. Unclear whether other customers had been contacted, so other primary cases and asymptomatic/mild cases potentially missed.<br>- Transmission before the 20 January cannot be ruled out, especially between the 2 cases who developed symptoms on 21-22 January.<br>- Very little information provided on the methods and results of the investigations, only conclusions. Not specified when the investigation was conducted. |

| Author, setting, period, objective | Participants, methods | Results relating to transmission routes | Results relating to transmission modifying factors | Study quality and key limitations |
|---|---|---|---|---|
| | | | | - Fomite transmission in bathrooms was not considered. |
| Katelaris et al, 2021 [8]<br>Setting: church singing in Sydney, Australia<br>Study period: Exposure 15-17 July 2020; public health authorities notified on 20 July, investigation started on 21 July 2020<br>Objective: to investigate the outbreak and assess the possibility of airborne transmission of SARS-CoV-2 | Participants: 1 primary case and 508 close contacts across the 4 church services (12 secondary cases).<br>Outcome assessment<br>- COVID-19 test (nasopharyngeal swab; RT-PCR) of all participants (85% uptake).<br>- Close contacts asked for symptoms every 2/3 days during quarantine<br>- Genomic sequencing for the primary case and 10 secondary cases.<br>Exposure assessment<br>- Interviews with cases<br>- Video recording<br>- Site visits with building managers to understand ventilation. | - Primary case: 18-year-old choir member who, following SARS-CoV-2 exposure on 11 July, reported symptom onset on 16-17 July. Sang at 4 1h church services: 15, 16, and twice on 17 July 2020.<br>- All attendees to the 4 services were considered close contacts (n=508) and required to self-isolate and be tested (85% uptake of testing). The first two cases had been notified on 20 July, and most contacts were tested 2-7 days after exposure.<br>- 12 secondary cases were identified (SAR 2.4%). All had attended services on 15 and/or 16 July, none had attended services only on 17 July.<br>- Video analysis found all secondary cases sat in the same section, 1-15m from the primary case, who was located in a choir loft 3.5m above the congregation, facing away from the secondary cases. No cases were detected in other sections of the church.<br>- Except for 5 of the secondary cases who were from the same household, close contact and fomite transmission unlikely as primary case denied mixing with attendees or touching objects (confirmed by video analysis). | Insufficient air replacement:<br>- Lack of ventilation and aeration (ventilation system and fans not in operation; windows and doors closed, except for entrance and exit).<br>Singing<br>May have increased the amount of aerosols generated by the primary case. | Quality rating: high<br>Key limitations<br>- Singing as a modifying factor for airborne transmission was presented as a hypothesis, no data presented to support it.<br>Testing performed within one week of exposure, some cases might have been missed. |

| Author, setting, period, objective | Participants, methods | Results relating to transmission routes | Results relating to transmission modifying factors | Study quality and key limitations |
|---|---|---|---|---|
| | | - Transmission outside the outbreak deemed unlikely as community transmission was low at the time and genome sequencing suggested single cluster. | | |
| Kwon et al, 2020 [9]<br><br>Setting: restaurant in Jeonju, Korea<br><br>Study period: case A visited restaurant on 12 June 2020, tested positive for COVID-19 on 17 June and epidemiological field investigation took place 19 June – 2 July 2020<br><br>Objective: to investigate how transmission of SARS-CoV-2 occurred in a restaurant | Participants: 14 participants, including 1 primary case (case B) and their 13 close contacts at the restaurant (11 visitors and 2 employees).<br><br>Outcome assessment<br>- COVID-19 test (nasopharyngeal swab, RT-PCR) for all close contacts<br>- Genome sequencing for positive cases<br><br>Exposure assessment<br>- Contact tracing<br>- Personal interviews<br>- Credit card records<br>- CCTV images<br>- Mobile phone location data<br>- On-site visits for environmental sampling (RT-PCR, 39 samples from air conditioning units, tables and chairs) and to assess restaurant structure, seating arrangement and air flow (anemometer) | - Case B (primary case) visited the restaurant on 12 June 2020, 1 day before symptom onset.<br>- 13 persons identified as close contacts, of which 2 tested positive: case A (symptom onset 16 June, positive test 17 June) and case B (symptom onset 18 June, positive test 20 June); link to primary case confirmed by genomic analysis. Secondary attack rate: 15.4% (2/13).<br>- Cases B and A: in the restaurant at the same time for 5min, 6.5m apart (without mask). Close contact and fomite transmission unlikely as they did not use the same door and case A did not leave their table (CCTV).<br>- Cases B and C: in the restaurant at the same time for 21min, 4.8m apart. Fomite transmission through door handle unlikely as they did not use the same door.<br>- All environmental samples tested negative (qRT-PCR). | Recirculating air flow<br>- Air conditioner units created air flow path from case B to cases A (maximum speed 1.0m/s) and C (maximum speed 1.2 m/s).<br>- The other visitors present in the restaurant (including some that were closer to case B and for a longer time) but not in the air flow path from case B did not get infected. Visitors sitting at tables with cases A and C but facing away from primary case did not get infected.<br><br>Insufficient air replacement<br>- No windows and no ventilation system. | Quality rating: high<br><br>Key limitations<br>- The authors ruled out close contact and fomite transmission; however they did not report results of CCTV analysis for interaction between cases B and C.<br>- One additional visitor tested positive for COVID-19 (case D, who was with case B at the restaurant) on 16 June (symptom onset 15 June). Not included in the investigation as believed to be part of a different cluster (supposedly infected on 11 June). However, no information provided on genomic analysis.<br>- Environmental sampling conducted on 23 June 2020, 11 days after event. |

| Author, setting, period, objective | Participants, methods | Results relating to transmission routes | Results relating to transmission modifying factors | Study quality and key limitations |
|---|---|---|---|---|
| Li et al, 2021[10]<br><br>(outbreak originally reported by Lu et al (2020)[11])<br><br>Setting: restaurant in Guangzhou, China<br><br>Study period: exposure 24 January 2020 (Chinese New Year's Eve). Tracer gas study, 19-20 March 2020<br><br>Objective: to investigate an outbreak involving 3 families that occurred at a restaurant and evaluate airborne transmission and associated environmental conditions | Participants: 89 visitors (10 positive cases) and 8 staff<br><br>Outcome assessment<br>- COVID-19 test (throat swab, RT-PCR) for all participants<br><br>Exposure assessment<br>- Epidemiological data (including travel and exposure history) and seating arrangement from Li et al[11]<br>- CCTV recording (of restaurant and elevator)<br>- Design of air conditioning and ventilation system<br>- Hourly weather data<br>- Computational fluid dynamics (CFD) simulation and tracer gas measurements (on-site visit) to study airflow and respiratory particles dispersion. | - 9 potential secondary and tertiary cases (symptom onset up to 6 Feb) at tables A, B and C situated at the back of the restaurant; potential primary case (symptom onset later on 24 Jan) sat at table A.<br><br>- Table A located between tables B and C; overlap time between tables A and B was 53min and 75min between tables A and C. Distance between primary case and potential secondary cases was 1.4m-4.6m.<br><br>- Table A: (visiting from Wuhan) 10 people from 4 households, 5 infected (including primary case); but transmission between primary and secondary cases could have happened outside restaurant.<br><br>- Table B: 4 people from 2 households, 3 infected. Table C : 7 people from 3 households, 2 infected. No contact with any known COVID-19 patients or visitors from Hubei Province 14 days prior to symptom onset. Unclear whether all had been infected at the restaurant, but likely that transmission happened at the restaurant for at least 1 member of each table.<br><br>- CCTV analysis: no risk of fomite transmission or close contact during lunch, in the toilet or in elevator (apart from some seating back to back); table A was active (standing up, speaking right and left, but primary case never turned their head towards table B) | Recirculating air flow by air circulation units<br>- 5 fan coil air conditioning units, one of which was at the back of the restaurant directed towards tables A, B and C.<br>- CFD simulation predicted a relatively isolated air recirculation zone around tables A B and C, which was supported by ethane gas experiments. Those on tables next to table A, but not in the circulating air stream, did not get infected.<br>- Experimentation showed higher gas concentrations associated with higher risk of being infected with COVID-19. (Odds ratio associated with a 1% increase in concentration: 1.115; 95% CI: 1.008–1.233; p=0.035).<br><br>Insufficient air replacement<br>- Air-conditioning units without outdoor air supply and exhaust fans not in use (except 1 in the bathroom providing occasional natural ventilation); door used approximately every 2 | Quality rating: medium<br><br>Key limitations<br>- Genomic sequencing not performed, transmission outside this event cannot be ruled out. Cases in table B had symptom onset between 1 and 5 Feb (≥8 days after event); could have been infected elsewhere.<br>- No risk of close contact or fomite transmission reported by the study authors, however all members of 3 tables used the bathroom. Whilst there was no overlap with the primary case, fomite transmission cannot be ruled out. |

| Author, setting, period, objective | Participants, methods | Results relating to transmission routes | Results relating to transmission modifying factors | Study quality and key limitations |
|---|---|---|---|---|
| | | while tables B and C were rather inactive in comparison. | minutes but no windows opened.<br>- 2 tracer gas decay experiments showed the air exchange rate was only 0.77 air changes/hour and ventilation rate 0.75–1.04 L/s fresh air/person. | |
| Lin et al, 2021 [12]<br><br>Setting: 29-storey apartment with 3 units in Guangzhou, China<br><br>Study period: primary case diagnosed with COVID-19 on 27 January 2020, unit evacuated on 8 February, investigation period not specified<br><br>Objective: to investigate a community outbreak in apartments and evaluate airborne transmission | Participants: 9 symptomatic cases from 3 flats in unit B; total number of residents not specified<br><br>Outcome assessment<br>- COVID-19 test (nasopharyngeal swab; RT-PCR) of the 9 symptomatic cases<br>- Genome sequencing.<br><br>Exposure assessment<br>- Interviews with cases<br>- CCTV of the elevator<br>- Simulated experiments within the apartments with tracer gas and measurement of air flow. | - All 9 cases were symptomatic and genomic analysis confirmed viral cluster. 5 cases from flat 15b (tested positive 26-29 January), 2 from 25b (tested positive 1 February) and 2 from 27b (tested positive 6-13 February).<br>- 4 of the 5 members of flat 15b had travel history to Wuhan; flats 25b and 27b did not.<br>- Elevator disinfected immediately after diagnosis of the primary case. Elevator CCTV showed no close contacts within the elevator between 25 and 27 January, and that family 15b wore masks every time but once when in the elevator; families 25b and 27b did not.<br>- Elevator used by residents of all 3 units. However, significant difference in chance of testing positive for residents from unit b compared to residents not from unit b ($p<0.05$; Fisher's exact test): location of positive cases was unlikely to be due to | Insufficient air replacement<br>- Ventilation efficiency lower in unit b than in unit a and c due to a modification of the ventilation pipe (narrower and bent at a right angle). Tracer gas experiment showed gas remained for longer in the pipes compared to unit A (>60min vs <30 min).<br>- Windows were closed (winter, cold weather).<br><br>Recirculating air flow:<br>- Wind speed experiment showed that on flushing a toilet, strong airflow could drive virus through drainage and exhaust system for vertical line transmission between connected apartments on different floors. | Quality rating: medium<br><br>Key limitations<br>- Only symptomatic testing, asymptomatic cases in others flats and units might have been missed.<br>- CCTV analysis of elevator limited to 25-27 January, close contact or fomite transmission before or after this period cannot be ruled out.<br>- Unclear whether possibility of close contacts in building entrance and corridors was considered.<br>- The authors reported in the discussion that residents generally wore masks outside of their apartments and avoided going outside, but it is unclear whether this started |

| Author, setting, period, objective | Participants, methods | Results relating to transmission routes | Results relating to transmission modifying factors | Study quality and key limitations |
|---|---|---|---|---|
| | | chance alone, or due to transmission in the elevator.<br>- Families 25b and 27b reported no close contacts with family 15b or with other cases.<br>- All cases located in unit b, sharing a common pipe system. Wind speed and tracer-gas experiments showed that long-distance airborne transmission through pipe system was possible. | | before or after the outbreak had been detected. |
| Luo et al, 2020 [13]<br>Setting: public transport (coach and minibus), Hunan province, China<br>Study period: bus trips on 22 January 2020, investigation period not specified<br>Objective: contact tracing study as part of an outbreak study | Participants: primary case and 243 potential contacts; 9 secondary cases identified who had travelled with primary case<br>Outcome assessment<br>- COVID-19 test (nasopharyngeal swab; RT-PCR) for all participants<br>Exposure assessment<br>- Epidemiological survey (travel history and close contacts of suspected cases)<br>- Bus seating layouts and loading and unloading stops of all passengers (obtained from public transportation authority). | - Primary case (symptom onset 22 January; tested positive 29 January) travelled on 2 buses on 22 January:<br>• First journey (coach, 2.5h): of the 48 passengers (including driver), 7 tested positive (symptom onset: 23 January - 4 February, 1 asymptomatic).<br>• Second journey (minibus, 1h): of the 12 passengers (including driver), 2 tested positive (symptom onset: 24 January and 31 January).<br>- Secondary attack rate for both journeys: 15%; 95% CI 6% to 24%.<br>- Majority of secondary cases >2m from primary case, up to 4.5m.<br>- None of the cases wore face coverings on the buses.<br>- Fomite transmission and close contact transmission cannot be ruled out, although | Insufficient air replacement<br>- All windows were closed on the coach and on the minibus. Ventilation systems were on.<br>Recirculating air flow from ventilation system<br>- The coach had an exhaust fan in the front and ventilation inlets on both sides, possibly creating an air flow from the rear of the coach (where the primary case was seated) to the front. The minibus had an exhaust fan in the centre. | Quality rating: medium<br>Key limitations<br>- Genomic sequencing not performed so transmission outside this event cannot be ruled out, especially for 3 of the secondary cases who had symptom onset or tested positive less than 2 days after the journey.<br>- Unclear when the epidemiological investigation and the testing of the contacts took place. |

| Author, setting, period, objective | Participants, methods | Results relating to transmission routes | Results relating to transmission modifying factors | Study quality and key limitations |
|---|---|---|---|---|
| | | deemed unlikely for at least some cases (e.g. who had used different doors and did not report direct contact with primary case).<br>- None of the infected passengers had been in contact with a COVID-19 case in the two weeks prior to symptom onset. Very few cases reported in the Hunan Province before 22 January. | | |
| Shah et al, 2021 [14]<br>**PREPRINT (v2, 6 July 2021)**<br>Setting: 5 singing events, Netherlands<br>Study period: September and October 2020<br>Objective: to investigate whether singing increased SARS-CoV-2 transmission risk during singing events | Participants: between 9 (event 5) and 21 (event 2); 78 in total<br>Outcome assessment<br>- COVID-19 test (respiratory sample; RT-PCR) for all symptomatic cases (and for 4 asymptomatic cases); probable cases based on symptoms only<br>- Genome sequencing for some of the cases from events 4 and 5.<br>Exposure assessment<br>- Phone/email conversations with spokesperson of each group, and questionnaire for all participants (81% response rate, ranging from 58% for event 1 to 100% for event 4), including data on possible exposure within and | - National recommendations for singing event at the time of the study included physical distancing and ventilation. Rooms ranged between 320 to 3,000m$^3$.<br>- Event 1: 90 min duration (50 min singing), 19 attendees, 14 confirmed cases (74% attack rate), no single primary case identified (7 had symptom onset in the 3 days following the event). Cases widely dispersed in the room (likely >2 metres). Some staff present in the venue but no information available.<br>- Event 2: 120 min duration (80 min singing), 21 attendees, 13 confirmed cases, 1 probable case (67% attack rate), 2 possible primary cases identified. Cases widely dispersed in the room (likely >2 metres).<br>- Event 3: 150 min duration (120 min singing), 15 attendees, 8 confirmed cases (53% attack rate). 1 possible primary case identified. Cases widely dispersed in the room (likely >2 metres). | Insufficient air replacement<br>- Doors or/and windows reported to be opened in all events. In addition: event 3: ceiling ventilation; event 4: Possible mechanical ventilation.<br>- Not enough information provided to assess exact air exchange rates, but estimated to be about 3 air exchanges per hour (ACH) for events 1 and 5, and <1 ACH for the other events.<br>Recirculating air flow<br>- Members in events 1, 3 and 4 reported feeling an air draft.<br>- Not enough information to assess whether air flow was a modifying factor, but cannot be ruled out: air flow could have been generated through | Quality rating: medium<br>Key limitations<br>- Epidemiological investigation mainly based on questionnaire and unclear how long after the events it was done (risk of recall bias).<br>- Genomic sequencing not performed for 3 events, and for only 2 out of 8 cases for another event.<br>- Potential primary cases identified though symptom onset date (in all events, at least 1 participant had symptom onset in the 3 days following the event); asymptomatic transmission was not considered (no asymptomatic testing). |

| Author, setting, period, objective | Participants, methods | Results relating to transmission routes | Results relating to transmission modifying factors | Study quality and key limitations |
|---|---|---|---|---|
| | outside the event, and seating arrangements<br>- National Notifiable Diseases Surveillance System data<br>- Aerosol transmission model (AirCoV2) | - Event 4: 120 min duration (90 min singing), 14 attendees, 7 confirmed cases, 1 probable (57% attack rate), 1 possible primary case identified. Cases widely dispersed in the room (likely >2 metres). Genome sequencing: 2 participants on opposite sides of room had identical strain.<br>- Event 5: 60 min duration (20 min singing), 9 participants, 6 confirmed cases (67% attack rate), 1 possible primary case identified. Cases were positioned up to 3m from suspected primary case. Genome sequencing: 4 of 5 identical strains in participants sitting near one another.<br>- Droplet transmission reported to be unlikely in events 2 and 5, but possible for some secondary cases in events 1, 3 and 4 (lack of social distance reported before, after or during the break, and some members travelled together to and from the events); 6 participants lived together, all tested positive.<br>- Fomite transmission reported to be unlikely in events 1, 2, 3 and 5. Event 4: cannot be ruled out due to use of a coffee machine with a push button.<br>- Most attendees did not report contact with confirmed cases before (n=3) or after (n=4) the event and only 3 of all attendees reported having participated in another | opened doors or windows; and all events except event 2 had some mechanical ventilation systems (events 3 and 4) or heating systems (events 1 and 5).<br>Singing<br>- Possible modifying factors in all 5 events.<br>- AirCoV2 model suggests high virus concentration (eg presence of a supershedder with $10^{10}$ virus/mL of mucus) required to explain high attack rates observed in these 5 events. | |

| Author, setting, period, objective | Participants, methods | Results relating to transmission routes | Results relating to transmission modifying factors | Study quality and key limitations |
|---|---|---|---|---|
| | | singing event in the 14 days prior to the event. | | |
| Shen et al, 2020 [15]<br><br>Setting: bus transport to an outdoor religious event in Zhejiang province, China<br><br>Study period: bus ride 19 January 2020. Epidemiological investigation 27 January – 23 February 2020<br><br>Objective: to investigate how transmission of SARS-CoV-2 occurred during bus travel (with nested case control study) | Participants: 300 individuals attended religious event, of which 128 had travelled by bus (60 on bus 1 and 68 on bus 2; the 2 buses had similar design).<br><br>Outcome assessment<br>- COVID-19 test (throat swab; RT-PCR) for those involved in the outbreak and their close contact<br>Exposure assessment<br>- Questionnaires and interviews (demographics, travel history, seating arrangement in bus, etc)<br>- Contact tracing data<br>- Bus design and ventilation system | - 31 cases including primary case (first to develop symptoms and had close contact 2 days before event with 4 individuals with travel history to Hubei). Primary case considered presymptomatic as reported first symptoms after the event although a follow-up investigation suggested that they had a mild cough from the day before the event.<br><br>- 23 secondary cases had travelled in the same bus (bus 2) as the primary case (2 x 50 min travel time). The remaining 7 did not travel in a bus but reported close contact with primary case during the religious event (150min duration; outdoor). No secondary cases identified within the 60 individuals who travelled in bus 1. None of the participants wore masks and no IPC measures in place.<br><br>- For the 23 secondary cases from bus 2, most transmission events likely to have happened during bus ride rather than during religious event as otherwise cases would have been randomly scattered between buses. The event included a 15-30min lunch with tables of 10 during which passengers from bus 2 were randomly mixed. Relative risk (RR) of bus 2 compared to:<br>• Bus 1: RR 42.2 (95%CI 2.6 to 679.3; p<0.01) | Insufficient air replacement<br>- Bus air conditioning system was in indoor recirculation mode (warm air); no information provided on whether the 4 windows were opened.<br><br>- The driver and passengers seated near the door were not infected. Only 1 participant seated near openable window infected.<br><br>Recirculating air flow<br>- 16 air vents across both sides of the bus, no information provided on whether these might have resulted in air flow. | Quality rating: medium<br><br>Key limitations<br>- All individuals “involved in the outbreak” likely to be only those who developed symptoms (and their close contacts) rather than all participants to the event.<br>- Genomic sequencing reported in methods, but results not presented. It is likely that the genomic sequencing was used as case definition (see supplementary material of the paper) rather than to ensure cases belonged to the same genomic cluster.<br>- Authors commented on repartition of cases compared to position near windows, but not enough information provided to assess its significance in relation to air replacement and air flow. |

| Author, setting, period, objective | Participants, methods | Results relating to transmission routes | Results relating to transmission modifying factors | Study quality and key limitations |
|---|---|---|---|---|
| | | • All participants but those in bus 2: RR 11.4 (95%CI 5.1 to 25.4; p<0.01)<br>- Cases scattered within bus 2, no statistically significant association with being seated <2m from primary case. Severity of cases not associated either with proximity to primary case. Passengers remained seated during the ride and had same seats in on both journeys.<br>- Fomite transmission e.g. from a pole on the bus cannot be ruled out. | | |
